# Neck pain service utilization and costs: association with timing of non-pharmaceutical services for individuals initially contacting a primary care provider. A retrospective cohort study

**DOI:** 10.1101/2023.01.10.23284193

**Authors:** David Elton, Meng Zhang

**Author notes:** **Corresponding author:** David Elton, Optum Labs, 11000 Optum Circle, Eden Prairie, MN 55344. **Declarations**. **Ethics approval and consent to participate** Because the data was de-identified or a Limited Data Set in compliance with the Health Insurance Portability and Accountability Act and customer requirements, the UnitedHealth Group Office of Human Research Affairs determined that this study was exempt from Institutional Review Board review. **Consent for publication** Not applicable. **Availability of data and materials** The data are proprietary and are not available for public use but, under certain conditions, may be made available to editors and their approved auditors under a data-use agreement to confirm the findings of the current study. **Funding** None. **Authors’ contributions** Study conception and design; **DE**. Data acquisition; **DE, MZ**. Data analysis and interpretation; **DE, MZ**. Draft or substantially revise manuscript; **DE, MZ**.

## Abstract

**Background:** Neck pain (NP) is prevalent and costly. NP clinical practice guidelines are similar to those for low back pain (LBP), emphasizing non-pharmaceutical and non-interventional first-line approaches. Primary care providers (PCP) are frequently consulted by individuals with NP.

**Objective:** Examine the association between guideline concordant incorporation of non-pharmaceutical therapies, use of imaging, pharmaceutical, and interventional services, and total episode cost for individuals with NP initially contacting a PCP.

**Design:** Retrospective cohort study using identical methods to a previous LBP study

**Setting/Patients:** National sample of individuals with non-surgical NP occurring in 2017-2019.

**Measurements:** Independent variables were initial contact with a PCP, and the timing of incorporation of 5 types of non-pharmaceutical therapies. Dependent measures included use of 13 types of health care services and total episode cost.

**Results:** 70,252 PCPs were initially contacted by 124,780 individuals with 137,274 episodes of non-surgical NP. 30.9% of PCPs and 22.1% of episodes included at least one of five non-pharmaceutical services at any time during an episode. Active care (13.7% of episodes), manual therapy (10.8%), and chiropractic manipulative therapy (9.4%) were the most common non-pharmaceutical services. 7.4% of episodes included a non-pharmaceutical service during the first 7 days with these episodes associated with a modest reduction (risk ratio 0.28 to 0.78) in the use of prescription pharmaceuticals. Younger individuals from ZIP codes with higher adjusted gross income were more likely to receive a non-pharmaceutical service in the first 7 days of an episode. When included during an episode, non-pharmaceutical services were associated with an increase in total episode cost with the smallest increase associated with chiropractic and osteopathic manipulation.

**Limitations:** As a retrospective observational analysis of associations there are numerous potential confounders and limitations.

**Conclusions:** Non-pharmaceutical services are infrequently provided to individuals with non-surgical NP initially contacting a PCP. For these individuals, non-pharmaceutical services, if provided, are most commonly introduced later in an episode after receiving pharmaceutical, imaging, and interventional services. For individuals with NP initially contacting a PCP there is an opportunity to increase the guideline concordant incorporation of non-pharmaceutical services early in an episode.

## Introduction

Neck pain (NP) is prevalent, costly, and associated with a high number of years lived with disability.^1-7^ NP is a common reason for a visit to a primary care physician (PCP).^8-10^ After chiropractors PCPs are the second most common type of health care provider (HCP) initially contacted by individuals with NP.^11^

Among spinal disorders, management of low back pain (LBP) benefits from the availability of relatively homogenous high-quality clinical practice guidelines (CPGs) that describe a stepped approach in which services are sequenced into first-, second- and third-line services.^12-14^ In the absence of red flags of serious pathology, LBP CPGs emphasize individual self-management, non-pharmaceutical, and non-interventional services as first-line approaches.^12-14^ NP CPGs, while less common and more heterogenous than CPGs for LBP, generally recommend a similar approach where non-pharmaceutical and non-interventional approaches are recommended as initial management options.^15-26^

Spinal disorders, particularly LBP, have been identified as a source of a high proportion of non-guideline concordant “low-value” care, described as services generating costs without or with minimal beneficial impact on outcomes.^27,28^ The time burden associated with providing CPG concordant care, including for NP, is considerable.^8,29,30^ Examples of low-value care for LBP include overuse of imaging, interventional procedures, and some prescription medications such as opioids.^31-34^ The lower prevalence of NP and heterogeneity of NP CPGs results in less being known about the magnitude of low-value care for NP.

The type of HCP initially contacted has been used as a method to evaluate variation in service utilization and cost outcomes for LBP and NP.^11,35-38^ When initially contacted by an individual with NP, PCPs generally incorporate imaging and pharmaceutical services more frequently than CPG recommended non-pharmaceutical therapies.^35^ Several barriers to PCP referral for non-pharmaceutical therapies have been identified for spinal disorders.^39-46^ These barriers include coverage limitations ^39,45^, inconvenient access ^40,45^, cost ^40,41,45^, lack of familiarity and communication ^42-44^, limited time to make a referral ^45^, and concerns about possible adverse events.^46^

The aim of this retrospective, observational study was to replicate the methods used in an earlier study of LBP ^47^ to examine the associations between frequency and timing of incorporation of active care (AC), manual therapy (MT), chiropractic manipulative therapy (CMT), osteopathic manipulative therapy (OMT), or acupuncture (Acu) services, utilization of other healthcare services, and total episode cost for individuals with non-surgical NP initially contacting a PCP. Like the previous LBP study ^47^ the hypothesis was that early incorporation of one or more non-pharmaceutical services would be associated with lower rates of imaging, pharmaceutical and interventional service use, and lower total episode cost.

## Methods

### Study design, population, setting and data sources

This was a retrospective cohort study of individuals initially contacting a PCP for non-surgical NP. The study design was identical to a previous study of LBP conducted by the same author group.^47^ De-identified enrollment records and administrative claims data for individuals with NP were included in an enrollee database. HCP de-identified demographic information and professional licensure status was included in a HCP database. ZIP code level adjusted gross income (AGI) data was extracted from the Internal Revenue Service ^48^, population race and ethnicity data from the US Census Bureau ^49^, and socioeconomic status (SES) Area Deprivation Index (ADI) data, from the University of Wisconsin Neighborhood Atlas^®^ database.^50^

With study data de-identified or a Limited Data Set in compliance with the Health Insurance Portability and Accountability Act (HIPAA) and customer requirements the UnitedHealth Group Office of Human Research Affairs determined that this study was exempt from Institutional Review Board review. The study was conducted and reported based on the Strengthening the Reporting of Observational Studies in Epidemiology (STROBE) guidelines.^51^ [Supplement – STROBE Checklist]

With the available data it was not possible to differentiate whether non-pharmaceutical services were provided by a HCP accessed directly by the individual with NP after initially contacting the PCP or provided following referral from the either PCP or another HCP. The standard practice of attempting to generate causal inferences by adjusting for measurable, yet incomplete, confounders such as age, sex and co-morbidities ^52,53^ using potentially inadequate approaches such as propensity score matching, ^54^ not only does not yield causal insights, the distorted results can potentially limit translation potential. As an alternative, in this observational study actual demographic and episodic measures and associations are reported to facilitate straightforward translation. Examples of unmeasurable and potentially important confounders include: nuanced clinical complexity of NP not captured in administrative data, anticipated potential out of pocket costs and individual willingness to pay for different services, individuals self-paying for non-pharmaceutical or other services, availability of HCPs offering non-pharmaceutical services convenient to an individual’s home, workplace or daily travel routes, individual preference for specific services or types of HCP including gender or racial concordance, recommendations from family or friends, and appointment availability within a PCP’s and individual’s timing expectations for HCPs meeting these and other criteria.^55^

### Cohort selection and unit of analysis

The cohort included individuals aged 18 years and older initially contacting a PCP for a complete episode of NP commencing and ending between 1/1/2017 and 12/31/2019. This timeframe was selected as the period before the influence of the COVID-19 epidemic on care patterns in early 2020. All individuals had continuous medical and pharmacy insurance coverage during the entire study period.

Administrative claims data were translated into the episode of care unit of analysis using the Symmetry^®^ Episode Treatment Groups^®^ (ETG^®^) and Episode Risk Groups^®^ (ERG^®^) version 9.5 methodologies and definitions. ETG^®^ and ERG^®^ have been reported as a valid measurement for comparison of HCPs based on cost of care^56^, with previous studies finding a low risk of misclassification bias associated with the episode of care unit of analysis.^11,35,47^

Complete episodes, defined as having at least 91-day pre- and 61-day post-episode clean periods, were included in the analysis. Incomplete episodes were excluded. Also excluded were NP episodes including a surgical procedure, or episodes associated with diagnoses of malignant and non-malignant neoplasms, fractures and other spinal trauma, infection, congenital deformities and scoliosis, autoimmune disorders, osteoporosis, and advanced arthritis. As an observational analysis of associations, these straightforward exclusions were made to address a potential study limitation of individuals with more complex NP conditions influencing the timing of incorporation of first-line non-pharmaceutical and non-interventional services. By removing more complex diagnoses the analysis was able to focus on less complicated NP.

### Variables

Data preprocessing, table generation, and initial analyses were performed using Python (*Python Language Reference, Version 3*.*7*.*5*., n.d.). A goodness of fit analysis was conducted using D’Agostino’s K-squared test. Non-normally distributed data are reported using the median and interquartile range (IQR).

The primary independent variables were initial contact with a PCP, and the timing of incorporation of AC, MT, CMT, OMT, or acupuncture (Acu) services. The following Current Procedural Terminology (CPT®) codes were used to identify non-pharmaceutical services: AC – 97110, 97112, 97530; MT – 97140; CMT – 98940 to 98942; OMT – 98925 to 98929; Acu– 97810, 97811, 97813, 97814. For NP, these are the most frequently provided non-pharmaceutical services recommended by CPGs and covered by commercial insurance.^35^ For this study passive therapies were excluded from the definition of non-pharmaceutical services. The timing of incorporation of AC, MT, CMT, OMT, or Acu services was based on the number of days after the initial visit with a PCP when a non-pharmaceutical service was first billed by any HCP.^11,35,47^

The PCP HCP category consisted of Family Practice, Internal Medicine, General Medicine, and OBGYN physician types, along with Nurse Practitioner and Physician Assistant HCPs. The study cohort was able to access all PCP HCP types directly without a referral.^11,35,47^

The primary dependent variable was the rate and timing of use of 13 types of health care services.^11,35,47^ Secondary dependent variables included the total cost of care for all reimbursed services provided by any HCP during an episode, the number of different HCPs seen during an episode, and episode duration measured in days. Total episode cost included costs associated with all services provided for an episode of NP, including those not specifically identified in the 13 categories used in the analyses. Costs for services for which an insurance claim was not submitted were not available. The episode duration was the number of days between the first and last date of service for each episode.

Risk (RR) ratios, and associated 95% confidence intervals (CI), were calculated for the timing of introduction of each non-pharmaceutical service type compared to a baseline of a specific non-pharmaceutical service not being provided. Due to odds ratios tendency to exaggerate risk in common outcomes, RR were reported as the measure more widely understood in associational analyses.^57^ Bivariate analyses were also performed comparing episode attributes associated with timing of introduction of each non-pharmaceutical service type. Like the RR calculation the bivariate analysis baseline was episodes that did not include a specific non-pharmaceutical service. Differences in the percent of episodes including a service was evaluated using Fisher’s Exact test (p value of .001), with measures reported using median and IQR evaluated using Mann Whitney U test (p value of .001).

### Role of Funding Source

None

## Results

The sample included 124,780 individuals, with a median age of 47 (Q1 37, Q3 55), and 61.0% females. These individuals were associated with 137,274 complete non-surgical NP episodes involving 70,252 unique PCPs. There were $79,712,281 in reimbursed health care expenditures with a median total cost per episode of $157 (Q1 $51, Q3 $495). The median pre-episode clean period was 634 days (Q1 423, Q3 858). The median number of days between sequential episodes was 209 (Q1 119, Q3 346). The median post-episode clean period was 432 days (Q1 264, Q3 684) [Table 1]. Individuals were from all 50 States and some U.S. territories. [Supplement – State].

**Table 1.**
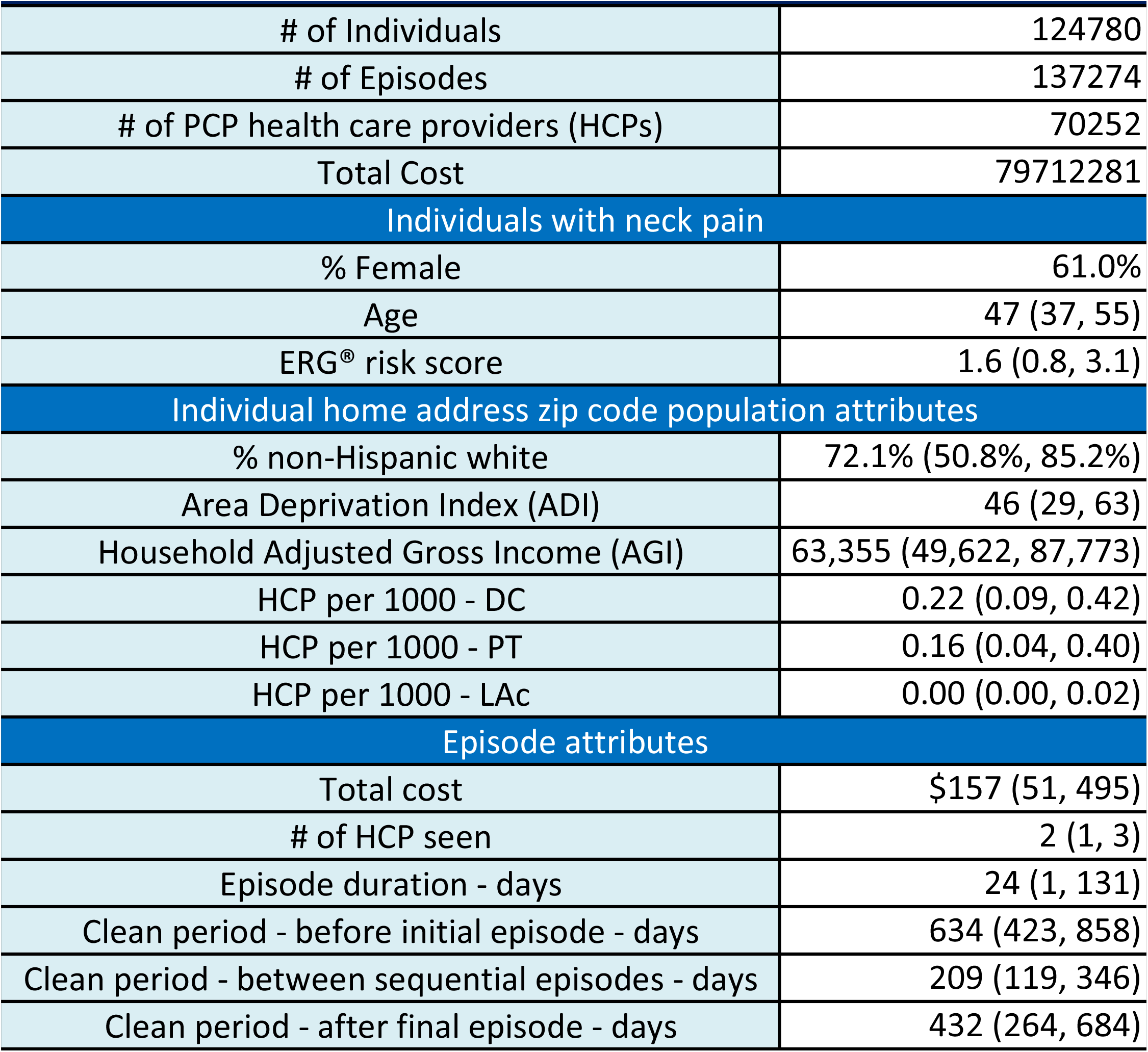
Cohort characteristics - % of Median (Q1, Q3)

|  |  |
| --- | --- |
| # of Individuals | 124780 |
| # of Episodes | 137274 |
| # of PCP health care providers (HCPs) | 70252 |
| Total Cost | 79712281 |
| Individuals with neck pain |  |
| % Female | 61.0% |
| Age | 47 (37, 55) |
| ERG® risk score | 1.6 (0.8, 3.1) |
| Individual home address zip code population attributes |  |
| % non-Hispanic white | 72.1% (50.8%, 85.2%) |
| Area Deprivation Index (ADI) | 46 (29, 63) |
| Household Adjusted Gross Income (AGI) | 63,355 (49,622, 87,773) |
| HCP per 1000 - DC | 0.22 (0.09, 0.42) |
| HCP per 1000 - PT | 0.16 (0.04, 0.40) |
| HCP per 1000 - LAc | 0.00 (0.00, 0.02) |
| Episode attributes |  |
| Total cost | \$157 (51, 495) |
| # of HCP seen | 2 (1, 3) |
| Episode duration - days | 24 (1, 131) |
| Clean period - before initial episode - days | 634 (423, 858) |
| Clean period - between sequential episodes - days | 209 (119, 346) |
| Clean period - after final episode - days | 432 (264, 684) |

77.9% of non-surgical NP episodes did not include a non-pharmaceutical service at any time during an episode. For the 22.1% of episodes that included any non-pharmaceutical service at any time during an episode, AC (13.7% of episodes), MT (10.8%) and CMT (9.4%) were most common. OMT (2.3% of episodes) and Acu (0.4%) were infrequently provided at any time during an episode. Individuals were more likely to receive skeletal muscle relaxants (34.8% of episodes), prescription NSAIDs (30.5%), radiography (25.8%), and opioids (15.7%) than non-pharmaceutical services. [Table 2]

**Table 2.**
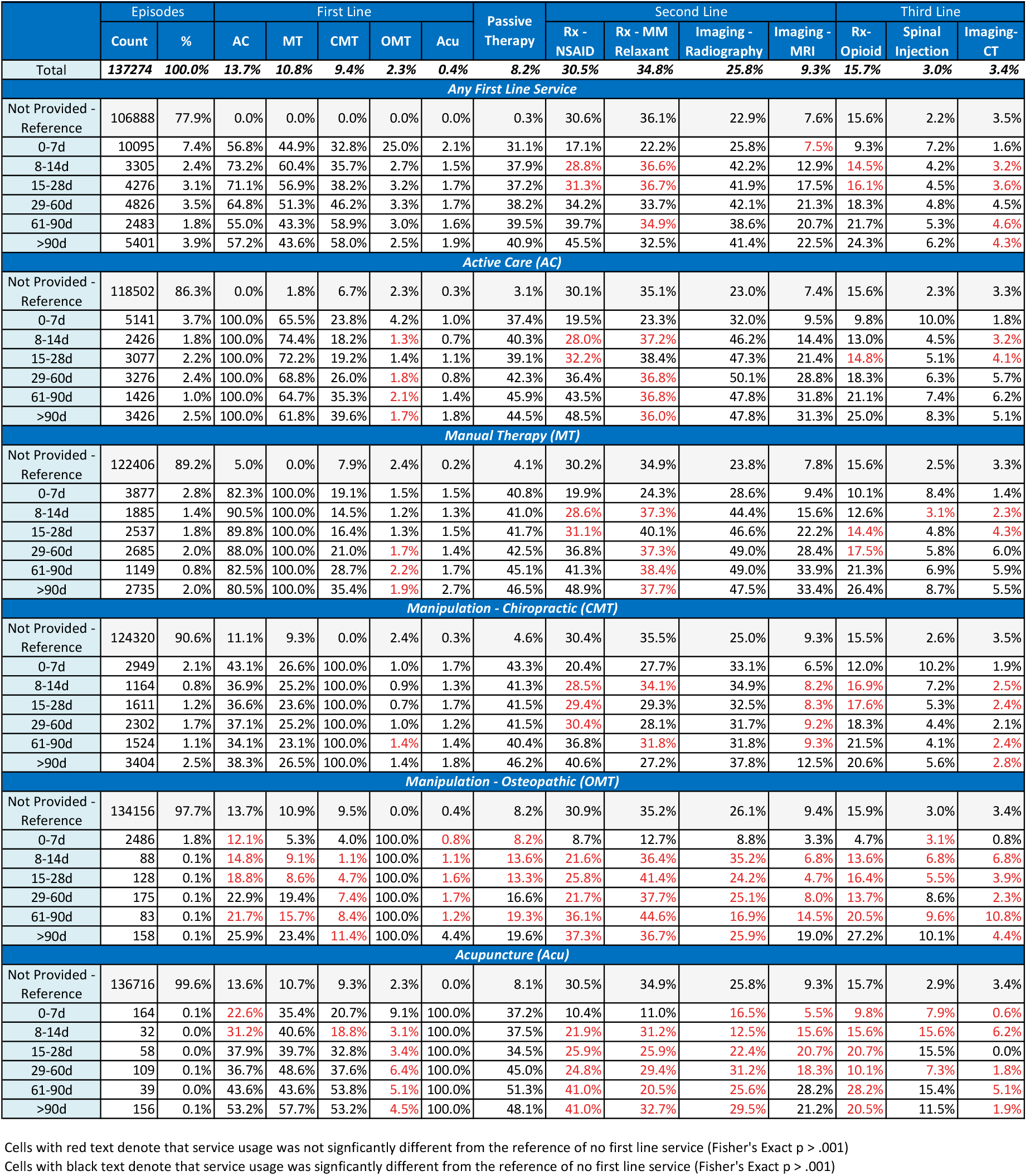
Non-surgical neck pain initially contacting PCP - episodic service use by number of days (d) into episode when first line service first incorporated.

|  |  |  |  |  |  |  |  |  |  |  |  |  |  |  |  |
| --- | --- | --- | --- | --- | --- | --- | --- | --- | --- | --- | --- | --- | --- | --- | --- |
|  | Episodes |  | First Line |  |  |  |  | Passive Therapy | Second Line |  |  |  | Third Line |  |  |
|  | Count | % | AC | MT | CMT | OMT | Acu |  | Rx - NSAID | Rx - MM Relaxant | Imaging - Radiography | Imaging - MRI | Rx- Opioid | Spinal Injection | Imaging- CT |
| Total | 137274 | 100.0% | 13.7% | 10.8% | 9.4% | 2.3% | 0.4% | 8.2% | 30.5% | 34.8% | 25.8% | 9.3% | 15.7% | 3.0% | 3.4% |
| Any First Line Service |  |  |  |  |  |  |  |  |  |  |  |  |  |  |  |
| Not Provided - Reference | 106888 | 77.9% | 0.0% | 0.0% | 0.0% | 0.0% | 0.0% | 0.3% | 30.6% | 36.1% | 22.9% | 7.6% | 15.6% | 2.2% | 3.5% |
| 0-7d | 10095 | 7.4% | 56.8% | 44.9% | 32.8% | 25.0% | 2.1% | 31.1% | 17.1% | 22.2% | 25.8% | 7.5% | 9.3% | 7.2% | 1.6% |
| 8-14d | 3305 | 2.4% | 73.2% | 60.4% | 35.7% | 2.7% | 1.5% | 37.9% | 28.8% | 36.6% | 42.2% | 12.9% | 14.5% | 4.2% | 3.2% |
| 15-28d | 4276 | 3.1% | 71.1% | 56.9% | 38.2% | 3.2% | 1.7% | 37.2% | 31.3% | 36.7% | 41.9% | 17.5% | 16.1% | 4.5% | 3.6% |
| 29-60d | 4826 | 3.5% | 64.8% | 51.3% | 46.2% | 3.3% | 1.7% | 38.2% | 34.2% | 33.7% | 42.1% | 21.3% | 18.3% | 4.8% | 4.5% |
| 61-90d | 2483 | 1.8% | 55.0% | 43.3% | 58.9% | 3.0% | 1.6% | 39.5% | 39.7% | 34.9% | 38.6% | 20.7% | 21.7% | 5.3% | 4.6% |
| >90d | 5401 | 3.9% | 57.2% | 43.6% | 58.0% | 2.5% | 1.9% | 40.9% | 45.5% | 32.5% | 41.4% | 22.5% | 24.3% | 6.2% | 4.3% |
| Active Care (AC) |  |  |  |  |  |  |  |  |  |  |  |  |  |  |  |
| Not Provided - Reference | 118502 | 86.3% | 0.0% | 1.8% | 6.7% | 2.3% | 0.3% | 3.1% | 30.1% | 35.1% | 23.0% | 7.4% | 15.6% | 2.3% | 3.3% |
| 0-7d | 5141 | 3.7% | 100.0% | 65.5% | 23.8% | 4.2% | 1.0% | 37.4% | 19.5% | 23.3% | 32.0% | 9.5% | 9.8% | 10.0% | 1.8% |
| 8-14d | 2426 | 1.8% | 100.0% | 74.4% | 18.2% | 1.3% | 0.7% | 40.3% | 28.0% | 37.2% | 46.2% | 14.4% | 13.0% | 4.5% | 3.2% |
| 15-28d | 3077 | 2.2% | 100.0% | 72.2% | 19.2% | 1.4% | 1.1% | 39.1% | 32.2% | 38.4% | 47.3% | 21.4% | 14.8% | 5.1% | 4.1% |
| 29-60d | 3276 | 2.4% | 100.0% | 68.8% | 26.0% | 1.8% | 0.8% | 42.3% | 36.4% | 36.8% | 50.1% | 28.8% | 18.3% | 6.3% | 5.7% |
| 61-90d | 1426 | 1.0% | 100.0% | 64.7% | 35.3% | 2.1% | 1.4% | 45.9% | 43.5% | 36.8% | 47.8% | 31.8% | 21.1% | 7.4% | 6.2% |
| >90d | 3426 | 2.5% | 100.0% | 61.8% | 39.6% | 1.7% | 1.8% | 44.5% | 48.5% | 36.0% | 47.8% | 31.3% | 25.0% | 8.3% | 5.1% |
| Manual Therapy (MT) |  |  |  |  |  |  |  |  |  |  |  |  |  |  |  |
| Not Provided - Reference | 122406 | 89.2% | 5.0% | 0.0% | 7.9% | 2.4% | 0.2% | 4.1% | 30.2% | 34.9% | 23.8% | 7.8% | 15.6% | 2.5% | 3.3% |
| 0-7d | 3877 | 2.8% | 82.3% | 100.0% | 19.1% | 1.5% | 1.5% | 40.8% | 19.9% | 24.3% | 28.6% | 9.4% | 10.1% | 8.4% | 1.4% |
| 8-14d | 1885 | 1.4% | 90.5% | 100.0% | 14.5% | 1.2% | 1.3% | 41.0% | 28.6% | 37.3% | 44.4% | 15.6% | 12.6% | 3.1% | 2.3% |
| 15-28d | 2537 | 1.8% | 89.8% | 100.0% | 16.4% | 1.3% | 1.5% | 41.7% | 31.1% | 40.1% | 46.6% | 22.2% | 14.4% | 4.8% | 4.3% |
| 29-60d | 2685 | 2.0% | 88.0% | 100.0% | 21.0% | 1.7% | 1.4% | 42.5% | 36.8% | 37.3% | 49.0% | 28.4% | 17.5% | 5.8% | 6.0% |
| 61-90d | 1149 | 0.8% | 82.5% | 100.0% | 28.7% | 2.2% | 1.7% | 45.1% | 41.3% | 38.4% | 49.0% | 33.9% | 21.3% | 6.9% | 5.9% |
| >90d | 2735 | 2.0% | 80.5% | 100.0% | 35.4% | 1.9% | 2.7% | 46.5% | 48.9% | 37.7% | 47.5% | 33.4% | 26.4% | 8.7% | 5.5% |
| Manipulation - Chiropractic (CMT) |  |  |  |  |  |  |  |  |  |  |  |  |  |  |  |
| Not Provided - Reference | 124320 | 90.6% | 11.1% | 9.3% | 0.0% | 2.4% | 0.3% | 4.6% | 30.4% | 35.5% | 25.0% | 9.3% | 15.5% | 2.6% | 3.5% |
| 0-7d | 2949 | 2.1% | 43.1% | 26.6% | 100.0% | 1.0% | 1.7% | 43.3% | 20.4% | 27.7% | 33.1% | 6.5% | 12.0% | 10.2% | 1.9% |
| 8-14d | 1164 | 0.8% | 36.9% | 25.2% | 100.0% | 0.9% | 1.3% | 41.3% | 28.5% | 34.1% | 34.9% | 8.2% | 16.9% | 7.2% | 2.5% |
| 15-28d | 1611 | 1.2% | 36.6% | 23.6% | 100.0% | 0.7% | 1.7% | 41.5% | 29.4% | 29.3% | 32.5% | 8.3% | 17.6% | 5.3% | 2.4% |
| 29-60d | 2302 | 1.7% | 37.1% | 25.2% | 100.0% | 1.0% | 1.2% | 41.5% | 30.4% | 28.1% | 31.7% | 9.2% | 18.3% | 4.4% | 2.1% |
| 61-90d | 1524 | 1.1% | 34.1% | 23.1% | 100.0% | 1.4% | 1.4% | 40.4% | 36.8% | 31.8% | 31.8% | 9.3% | 21.5% | 4.1% | 2.4% |
| >90d | 3404 | 2.5% | 38.3% | 26.5% | 100.0% | 1.4% | 1.8% | 46.2% | 40.6% | 27.2% | 37.8% | 12.5% | 20.6% | 5.6% | 2.8% |
| Manipulation - Osteopathic (OMT) |  |  |  |  |  |  |  |  |  |  |  |  |  |  |  |
| Not Provided - Reference | 134156 | 97.7% | 13.7% | 10.9% | 9.5% | 0.0% | 0.4% | 8.2% | 30.9% | 35.2% | 26.1% | 9.4% | 15.9% | 3.0% | 3.4% |
| 0-7d | 2486 | 1.8% | 12.1% | 5.3% | 4.0% | 100.0% | 0.8% | 8.2% | 8.7% | 12.7% | 8.8% | 3.3% | 4.7% | 3.1% | 0.8% |
| 8-14d | 88 | 0.1% | 14.8% | 9.1% | 1.1% | 100.0% | 1.1% | 13.6% | 21.6% | 36.4% | 35.2% | 6.8% | 13.6% | 6.8% | 6.8% |
| 15-28d | 128 | 0.1% | 18.8% | 8.6% | 4.7% | 100.0% | 1.6% | 13.3% | 25.8% | 41.4% | 24.2% | 4.7% | 16.4% | 5.5% | 3.9% |
| 29-60d | 175 | 0.1% | 22.9% | 19.4% | 7.4% | 100.0% | 1.7% | 16.6% | 21.7% | 37.7% | 25.1% | 8.0% | 13.7% | 8.6% | 2.3% |
| 61-90d | 83 | 0.1% | 21.7% | 15.7% | 8.4% | 100.0% | 1.2% | 19.3% | 36.1% | 44.6% | 16.9% | 14.5% | 20.5% | 9.6% | 10.8% |
| >90d | 158 | 0.1% | 25.9% | 23.4% | 11.4% | 100.0% | 4.4% | 19.6% | 37.3% | 36.7% | 25.9% | 19.0% | 27.2% | 10.1% | 4.4% |
| Acupuncture (Acu) |  |  |  |  |  |  |  |  |  |  |  |  |  |  |  |
| Not Provided - Reference | 136716 | 99.6% | 13.6% | 10.7% | 9.3% | 2.3% | 0.0% | 8.1% | 30.5% | 34.9% | 25.8% | 9.3% | 15.7% | 2.9% | 3.4% |
| 0-7d | 164 | 0.1% | 22.6% | 35.4% | 20.7% | 9.1% | 100.0% | 37.2% | 10.4% | 11.0% | 16.5% | 5.5% | 9.8% | 7.9% | 0.6% |
| 8-14d | 32 | 0.0% | 31.2% | 40.6% | 18.8% | 3.1% | 100.0% | 37.5% | 21.9% | 31.2% | 12.5% | 15.6% | 15.6% | 15.6% | 6.2% |
| 15-28d | 58 | 0.0% | 37.9% | 39.7% | 32.8% | 3.4% | 100.0% | 34.5% | 25.9% | 25.9% | 22.4% | 20.7% | 20.7% | 15.5% | 0.0% |
| 29-60d | 109 | 0.1% | 36.7% | 48.6% | 37.6% | 6.4% | 100.0% | 45.0% | 24.8% | 29.4% | 31.2% | 18.3% | 10.1% | 7.3% | 1.8% |
| 61-90d | 39 | 0.0% | 43.6% | 43.6% | 53.8% | 5.1% | 100.0% | 51.3% | 41.0% | 20.5% | 25.6% | 28.2% | 28.2% | 15.4% | 5.1% |
| >90d | 156 | 0.1% | 53.2% | 57.7% | 53.2% | 4.5% | 100.0% | 48.1% | 41.0% | 32.7% | 29.5% | 21.2% | 20.5% | 11.5% | 1.9% |

Cells with red text denote that service usage was not significantly different from the reference of no first line service (Fisher's Exact p > .001)
Cells with black text denote that service usage was significantly different from the reference of no first line service (Fisher's Exact p > .001)

Within the first 7 days of initially contacting a PCP 7.4% of episodes of non-surgical NP included one or more of the five non-pharmaceutical services with AC (3.7% of episodes), MT (2.8%) and CMT (2.1%) being most common. When introduced in the first 7 days after initial contact with a PCP, non-pharmaceutical services were generally associated with a reduction in exposure to prescription pharmaceuticals and an increase in exposure to radiology, MRI, and spinal injection services. OMT was associated with the largest reduction in exposure to prescription opioids (RR 0.29, 95% CI 0.25-0.25) followed by acupuncture (0.62, 0.39-.099), AC (0.63, 0.58-0.69), MT (0.65, 0.59-0.71) and CMT (0.77, 0.70-0.85). A non-pharmaceutical service introduced 8-14 days after initial contact with a PCP was associated with less significant and generally not clinically meaningful reduction in exposure to prescription NSAIDs and opioids along with an increase in exposure to spinal imaging and injections. When a non-pharmaceutical service was introduced 15+ days after initial contact with a PCP exposure to prescription pharmaceutical, spinal imaging and spinal injections were generally higher than if a non-pharmaceutical service was never provided [Table 2] [Supplement – Risk Ratio]. The RR for exposure to second- and third-line services based on timing of introduction of AC is illustrated in Figure 1, and CMT in Figure 2.

**Figure 1.**
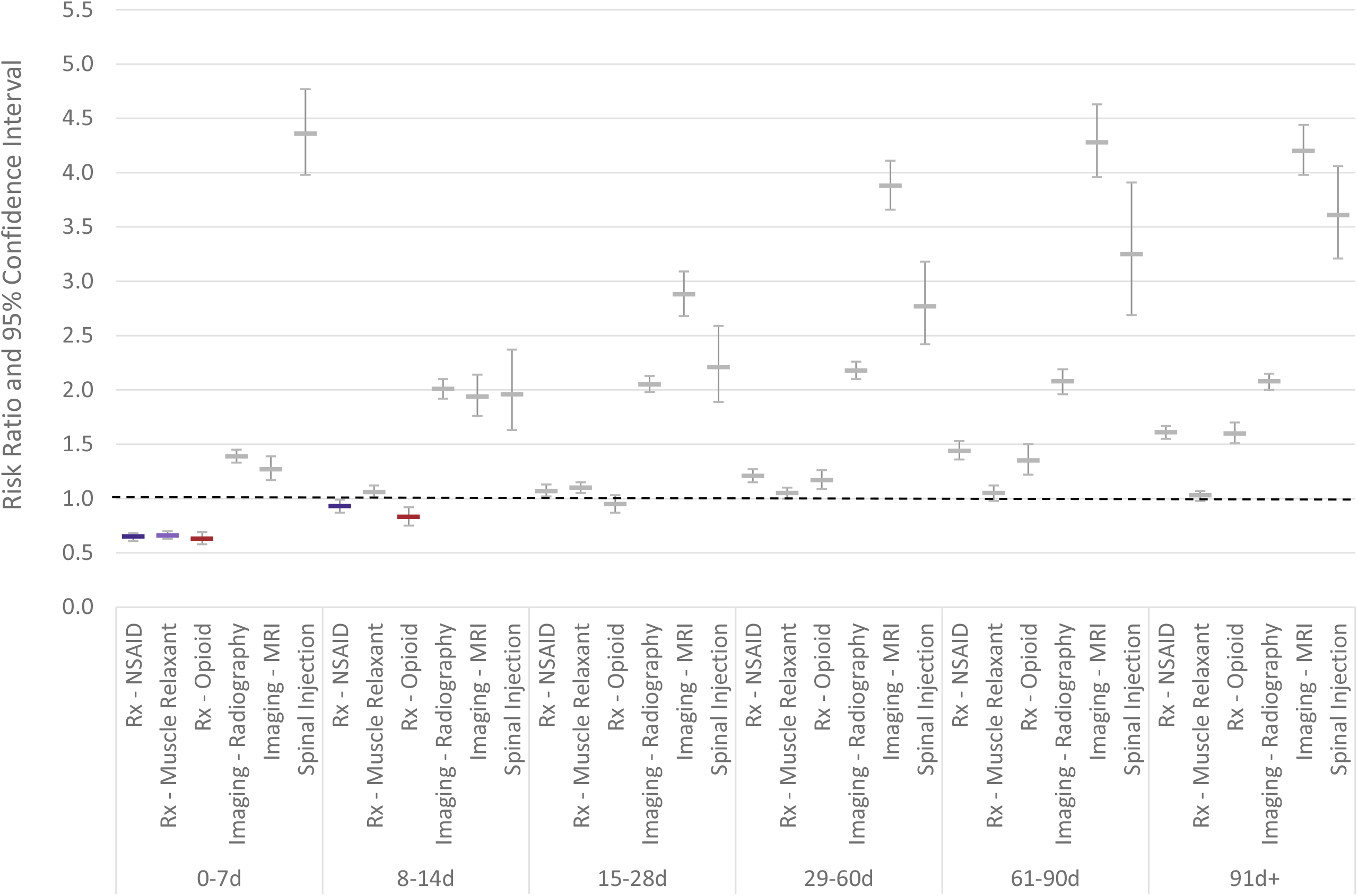
Individuals with non-surgical neck pain initially contacting a primary care provider. Risk ratio and 95% confidence interval for exposure to various health care services based on timing of introduction of **active care** compared to episodes without active care.

**Figure 2.**
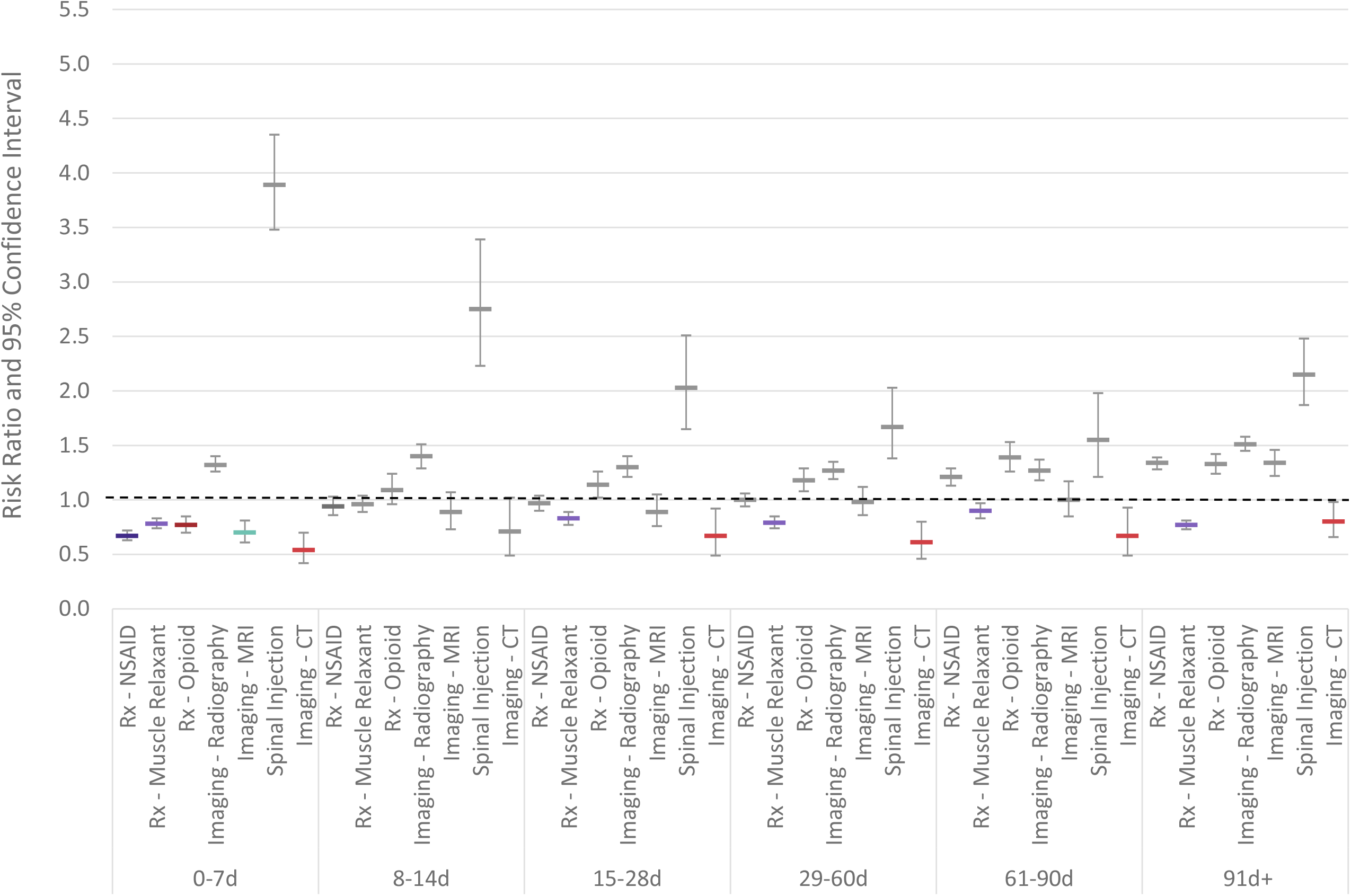
Individuals with non-surgical neck pain initially contacting a primary care provider. Risk ratio and 95% confidence interval for exposure to various health care services based on timing of introduction of **chiropractic manipulative therapy (CMT)** compared to episodes without CMT.

Episodes with a non-pharmaceutical service introduced within 7 days of initial contact with a PCP were associated with younger individuals, with a lower ERG^®^ risk score, from zip codes with lower deprivation, higher AGI, and greater availability of a chiropractor (DC) or physical therapist (PT). Among individual non-pharmaceutical services, Acu was most strongly associated with lower deprivation, higher AGI, lower percent non-Hispanic white population, and greater availability of a licensed acupuncturist (LAc) [Figure 3] [Table 3].

**Table 3.** Individual, local population and episode attributes associated with individuals with neck pain initially contacting a Primare Care Provider (PCP) by timing of incorporation of first line services.

|  |  |  |  |  |  |  |  |  |  |  |  |  |  |  |
| --- | --- | --- | --- | --- | --- | --- | --- | --- | --- | --- | --- | --- | --- | --- |
| % or Median (Q1, Q3) | PCPs |  | Episodes |  | Episode Attributes |  | Individuals |  | Individual Home Address Zip Code Population Attributes |  |  |  |  |  |
|  | Count | % | Count | % | Total Cost | Duration - days | Age | ERG® Risk | % non-Hispanic white (NHW) | ADI | AGI | HCP per 1000 Population |  |  |
|  |  |  |  |  |  |  |  |  |  |  |  | DC | PT | LAc |
| Total | 70252 | 100.0% | 137274 | 100.0% | \$157 (51, 495) | 24 (1, 131) | 47 (37, 55) | 1.6 (0.8, 3.1) | 72.1% (50.8%, 85.2%) | 46 (29, 63) | 63355 (49622, 87773) | 0.22 (0.09, 0.42) | 0.16 (0.04, 0.40) | 0.00 (0.00, 0.02) |
| Any First Line Service |  |  |  |  |  |  |  |  |  |  |  |  |  |  |
| Not Provided - reference | 48520 | 69.1% | 106888 | 77.9% | \$117 (30, 276) | 3 (1, 99) | 47 (37, 55) | 1.6 (0.8, 3.1) | 71.3% (49.5%, 84.8%) | 47 (30, 64) | 62358 (48946, 85692) | 0.21 (0.09, 0.41) | 0.16 (0.03, 0.39) | 0.00 (0.00, 0.02) |
| 0-7d | 6236 | 8.9% | 10095 | 7.4% | \$440 (170, 1088) | 35 (6, 112) | 44 (35, 54) | 1.5 (0.7, 2.8) | 73.7% (56.0%, 85.2%) | 42 (25, 58) | 69354 (53487, 99236) | 0.27 (0.12, 0.50) | 0.21 (0.06, 0.45) | 0.00 (0.00, 0.03) |
| 8-14d | 3134 | 4.5% | 3305 | 2.4% | \$721 (363, 1413) | 49 (27, 120) | 47 (37, 55) | 1.5 (0.8, 2.8) | 74.8% (57.2%, 86.6%) | 43 (27, 59) | 67505 (52271, 93975) | 0.25 (0.11, 0.47) | 0.20 (0.05, 0.47) | 0.00 (0.00, 0.03) |
| 15-28d | 4079 | 5.8% | 4276 | 3.1% | \$740 (333, 1560) | 63 (36, 144) | 47 (37, 55) | 1.6 (0.9, 3.0) | 74.0% (55.0%, 86.4%) | 43 (27, 59) | 67817 (52648, 93817) | 0.25 (0.11, 0.46) | 0.19 (0.05, 0.44) | 0.00 (0.00, 0.03) |
| 29-60d | 4576 | 6.5% | 4826 | 3.5% | \$732 (282, 1630) | 92 (57, 187) | 47 (38, 55) | 1.8 (1.0, 3.3) | 74.8% (55.8%, 87.0%) | 44 (27, 61) | 66500 (52057, 91574) | 0.25 (0.11, 0.47) | 0.18 (0.04, 0.44) | 0.00 (0.00, 0.03) |
| 61-90d | 2424 | 3.5% | 2483 | 1.8% | \$644 (226, 1562) | 133 (90, 229) | 47 (38, 55) | 2.0 (1.1, 3.7) | 75.9% (56.8%, 87.8%) | 46 (30, 62) | 64172 (51298, 85522) | 0.24 (0.11, 0.45) | 0.18 (0.04, 0.42) | 0.00 (0.00, 0.02) |
| > 90d | 5132 | 7.3% | 5401 | 3.9% | \$711 (280, 1724) | 232 (171, 317) | 48 (39, 56) | 2.2 (1.2, 3.8) | 76.4% (58.1%, 87.6%) | 46 (29, 62) | 64662 (51271, 89531) | 0.24 (0.11, 0.45) | 0.18 (0.05, 0.42) | 0.00 (0.00, 0.02) |
| Active Care |  |  |  |  |  |  |  |  |  |  |  |  |  |  |
| Not Provided - reference | 55891 | 79.6% | 118502 | 86.3% | \$127 (36, 310) | 9 (1, 114) | 47 (36, 55) | 1.6 (0.8, 3.1) | 72.1% (50.5%, 85.2%) | 47 (30, 64) | 62491 (49174, 85596) | 0.22 (0.09, 0.41) | 0.16 (0.04, 0.39) | 0.00 (0.00, 0.02) |
| 0-7d | 3371 | 4.8% | 5141 | 3.7% | \$728 (343, 1547) | 43 (15, 121) | 44 (35, 54) | 1.5 (0.8, 2.9) | 71.5% (52.3%, 82.6%) | 38 (23, 55) | 74134 (55276, 106439) | 0.26 (0.12, 0.49) | 0.21 (0.07, 0.45) | 0.00 (0.00, 0.04) |
| 8-14d | 2285 | 3.3% | 2426 | 1.8% | \$878 (498, 1658) | 47 (29, 106) | 47 (37, 55) | 1.4 (0.7, 2.7) | 73.2% (54.7%, 85.6%) | 40 (25, 57) | 69399 (53580, 101015) | 0.24 (0.10, 0.47) | 0.19 (0.06, 0.45) | 0.00 (0.00, 0.03) |
| 15-28d | 2932 | 4.2% | 3077 | 2.2% | \$960 (498, 1902) | 58 (37, 123) | 47 (37, 55) | 1.5 (0.8, 2.9) | 72.4% (52.9%, 84.6%) | 41 (25, 57) | 69722 (52970, 98984) | 0.24 (0.11, 0.45) | 0.20 (0.06, 0.46) | 0.00 (0.00, 0.03) |
| 29-60d | 3138 | 4.5% | 3276 | 2.4% | \$1076 (513, 2124) | 87 (57, 163) | 48 (38, 56) | 1.8 (0.9, 3.3) | 72.6% (52.7%, 84.6%) | 41 (25, 59) | 69200 (52824, 96876) | 0.23 (0.10, 0.45) | 0.18 (0.04, 0.45) | 0.00 (0.00, 0.03) |
| 61-90d | 1408 | 2.0% | 1426 | 1.0% | \$1139 (545, 2282) | 130 (93, 219) | 48 (39, 56) | 2.1 (1.1, 3.7) | 72.5% (52.9%, 85.9%) | 43 (26, 59) | 67737 (52133, 93673) | 0.23 (0.10, 0.43) | 0.19 (0.04, 0.44) | 0.00 (0.00, 0.03) |
| > 90d | 3295 | 4.7% | 3426 | 2.5% | \$1172 (560, 2369) | 233 (173, 320) | 49 (40, 56) | 2.4 (1.3, 4.1) | 74.4% (55.3%, 86.4%) | 43 (26, 60) | 67776 (52124, 95516) | 0.24 (0.11, 0.43) | 0.19 (0.05, 0.43) | 0.00 (0.00, 0.03) |
| Manual Therapy |  |  |  |  |  |  |  |  |  |  |  |  |  |  |
| Not Provided - reference | 58386 | 83.1% | 122406 | 89.2% | \$133 (39, 342) | 12 (1, 118) | 47 (36, 55) | 1.6 (0.8, 3.1) | 72.0% (50.5%, 85.2%) | 47 (30, 64) | 62612 (49287, 85969) | 0.22 (0.09, 0.42) | 0.16 (0.04, 0.39) | 0.00 (0.00, 0.02) |
| 0-7d | 2624 | 3.7% | 3877 | 2.8% | \$724 (323, 1523) | 43 (15, 120) | 45 (35, 54) | 1.5 (0.8, 2.9) | 72.1% (52.9%, 83.6%) | 37 (22, 55) | 75107 (55422, 108002) | 0.26 (0.12, 0.50) | 0.21 (0.06, 0.45) | 0.00 (0.00, 0.03) |
| 8-14d | 1825 | 2.6% | 1885 | 1.4% | \$897 (501, 1698) | 46 (29, 105) | 47 (37, 55) | 1.4 (0.7, 2.6) | 73.6% (56.5%, 85.9%) | 39 (24, 55) | 69676 (54246, 101098) | 0.25 (0.11, 0.47) | 0.21 (0.06, 0.48) | 0.00 (0.00, 0.03) |
| 15-28d | 2435 | 3.5% | 2537 | 1.8% | \$1031 (539, 2007) | 60 (38, 131) | 47 (37, 55) | 1.5 (0.8, 2.9) | 72.5% (53.0%, 84.7%) | 40 (24, 57) | 70070 (52990, 102037) | 0.25 (0.11, 0.45) | 0.20 (0.06, 0.46) | 0.00 (0.00, 0.04) |
| 29-60d | 2583 | 3.7% | 2685 | 2.0% | \$1132 (553, 2206) | 87 (57, 163) | 48 (39, 56) | 1.8 (1.0, 3.3) | 72.9% (54.0%, 85.1%) | 41 (24, 59) | 69545 (52620, 98682) | 0.24 (0.11, 0.46) | 0.19 (0.05, 0.47) | 0.00 (0.00, 0.03) |
| 61-90d | 1140 | 1.6% | 1149 | 0.8% | \$1218 (597, 2358) | 133 (92, 219) | 49 (39, 56) | 2.1 (1.2, 3.7) | 73.1% (53.2%, 85.7%) | 43 (26, 59) | 66782 (51672, 91814) | 0.23 (0.10, 0.44) | 0.19 (0.04, 0.44) | 0.00 (0.00, 0.03) |
| > 90d | 2667 | 3.8% | 2735 | 2.0% | \$1311 (605, 2557) | 234 (173, 323) | 49 (40, 56) | 2.4 (1.3, 4.0) | 74.7% (55.4%, 86.5%) | 42 (26, 60) | 68065 (52791, 95950) | 0.24 (0.11, 0.43) | 0.19 (0.05, 0.44) | 0.00 (0.00, 0.03) |
| Manipulation - Chiropractic |  |  |  |  |  |  |  |  |  |  |  |  |  |  |
| Not Provided - reference | 59507 | 84.7% | 124320 | 90.6% | \$142 (42, 428) | 14 (1, 111) | 47 (37, 55) | 1.6 (0.8, 3.1) | 71.5% (49.9%, 84.8%) | 46 (29, 63) | 63178 (49378, 87756) | 0.22 (0.09, 0.41) | 0.16 (0.04, 0.40) | 0.00 (0.00, 0.02) |
| 0-7d | 2177 | 3.1% | 2949 | 2.1% | \$418 (193, 1012) | 50 (10, 158) | 43 (34, 53) | 1.4 (0.8, 2.8) | 76.9% (59.5%, 86.9%) | 43 (27, 60) | 67876 (52998, 94477) | 0.28 (0.13, 0.51) | 0.20 (0.06, 0.47) | 0.00 (0.00, 0.03) |
| 8-14d | 1126 | 1.6% | 1164 | 0.8% | \$444 (188, 1015) | 69 (25, 172) | 45 (35, 54) | 1.6 (0.9, 3.1) | 77.3% (61.4%, 88.8%) | 48 (31, 63) | 64211 (50763, 85640) | 0.26 (0.12, 0.48) | 0.20 (0.05, 0.47) | 0.00 (0.00, 0.02) |
| 15-28d | 1572 | 2.2% | 1611 | 1.2% | \$436 (185, 979) | 85 (36, 197) | 47 (36, 55) | 1.8 (1.0, 3.3) | 77.3% (59.9%, 88.8%) | 46 (31, 61) | 65015 (52352, 87732) | 0.27 (0.12, 0.47) | 0.19 (0.05, 0.42) | 0.00 (0.00, 0.02) |
| 29-60d | 2226 | 3.2% | 2302 | 1.7% | \$400 (166, 985) | 112 (59, 224) | 46 (37, 54) | 1.8 (1.0, 3.2) | 77.6% (60.1%, 88.6%) | 46 (31, 63) | 64492 (51739, 87163) | 0.27 (0.12, 0.50) | 0.17 (0.05, 0.42) | 0.00 (0.00, 0.02) |
| 61-90d | 1496 | 2.1% | 1524 | 1.1% | \$390 (149, 958) | 140 (89, 241) | 46 (36, 54) | 1.9 (1.0, 3.5) | 77.9% (61.4%, 88.9%) | 47 (32, 62) | 63635 (51693, 84571) | 0.27 (0.12, 0.47) | 0.18 (0.04, 0.41) | 0.00 (0.00, 0.00) |
| > 90d | 3265 | 4.6% | 3404 | 2.5% | \$466 (205, 1151) | 239 (176, 323) | 47 (37, 55) | 2.0 (1.2, 3.4) | 77.3% (60.1%, 88.2%) | 47 (31, 62) | 64406 (51329, 87822) | 0.26 (0.11, 0.48) | 0.18 (0.05, 0.42) | 0.00 (0.00, 0.01) |
| Manipulation - Osteopathic |  |  |  |  |  |  |  |  |  |  |  |  |  |  |
| Not Provided - reference | 68689 | 97.8% | 134156 | 97.7% | \$156 (50, 494) | 24 (1, 132) | 47 (37, 55) | 1.6 (0.8, 3.1) | 72.0% (50.6%, 85.1%) | 46 (29, 63) | 63338 (49591, 87756) | 0.22 (0.09, 0.42) | 0.16 (0.04, 0.40) | 0.00 (0.00, 0.02) |
| 0-7d | 1175 | 1.7% | 2486 | 1.8% | \$157 (70, 360) | 2 (1, 65) | 44 (34, 53) | 1.4 (0.7, 2.6) | 76.2% (60.3%, 86.8%) | 45 (28, 61) | 64392 (51454, 92781) | 0.28 (0.13, 0.49) | 0.21 (0.05, 0.44) | 0.00 (0.00, 0.04) |
| 8-14d | 83 | 0.1% | 88 | 0.1% | \$446 (275, 660) | 36 (14, 136) | 47 (37, 55) | 1.2 (0.8, 2.8) | 76.8% (58.3%, 84.4%) | 42 (32, 59) | 66044 (51460, 86267) | 0.20 (0.09, 0.41) | 0.17 (0.05, 0.42) | 0.00 (0.00, 0.04) |
| 15-28d | 128 | 0.2% | 128 | 0.1% | \$366 (192, 931) | 51 (26, 153) | 40 (33, 52) | 1.6 (1.0, 3.5) | 71.2% (52.5%, 84.5%) | 48 (32, 61) | 62963 (51397, 80919) | 0.23 (0.11, 0.42) | 0.16 (0.03, 0.37) | 0.00 (0.00, 0.04) |
| 29-60d | 163 | 0.2% | 175 | 0.1% | \$445 (219, 1150) | 80 (53, 191) | 46 (37, 53) | 1.7 (1.0, 3.1) | 72.6% (56.6%, 83.8%) | 47 (30, 64) | 62035 (51073, 89515) | 0.29 (0.14, 0.44) | 0.19 (0.03, 0.35) | 0.00 (0.00, 0.03) |
| 61-90d | 81 | 0.1% | 83 | 0.1% | \$332 (153, 1489) | 134 (86, 190) | 46 (37, 54) | 2.2 (1.0, 4.0) | 75.3% (55.9%, 84.1%) | 47 (32, 64) | 66818 (51956, 83646) | 0.25 (0.09, 0.47) | 0.21 (0.05, 0.51) | 0.00 (0.00, 0.02) |
| > 90d | 152 | 0.2% | 158 | 0.1% | \$675 (299, 1786) | 246 (183, 342) | 48 (39, 57) | 2.3 (1.3, 4.0) | 77.7% (62.5%, 87.2%) | 45 (28, 63) | 64308 (50505, 84938) | 0.25 (0.14, 0.45) | 0.17 (0.04, 0.49) | 0.00 (0.00, 0.03) |
| Acupuncture |  |  |  |  |  |  |  |  |  |  |  |  |  |  |
| Not Provided - reference | 69783 | 99.3% | 136716 | 99.6% | \$156 (51, 490) | 23 (1, 131) | 47 (37, 55) | 1.6 (0.8, 3.1) | 72.2% (50.8%, 85.2%) | 46 (29, 63) | 63302 (49593, 87726) | 0.22 (0.09, 0.42) | 0.16 (0.04, 0.40) | 0.00 (0.00, 0.02) |
| 0-7d | 109 | 0.2% | 164 | 0.1% | \$376 (183, 915) | 30 (7, 95) | 45 (37, 53) | 1.4 (0.7, 2.6) | 66.4% (51.3%, 79.5%) | 20 (11, 35) | 94015 (72056, 134400) | 0.29 (0.15, 0.52) | 0.30 (0.10, 0.63) | 0.04 (0.00, 0.11) |
| 8-14d | 32 | 0.0% | 32 | 0.0% | \$628 (308, 1552) | 43 (23, 77) | 46 (40, 53) | 1.9 (0.9, 2.9) | 65.4% (46.2%, 78.5%) | 29 (17, 54) | 72939 (49691, 97044) | 0.32 (0.14, 0.46) | 0.26 (0.10, 0.54) | 0.03 (0.00, 0.08) |
| 15-28d | 56 | 0.1% | 58 | 0.0% | \$889 (287, 2024) | 65 (34, 222) | 46 (39, 52) | 1.9 (0.9, 3.9) | 66.1% (46.8%, 76.6%) | 31 (14, 46) | 85459 (63381, 111573) | 0.23 (0.13, 0.45) | 0.19 (0.08, 0.40) | 0.00 (0.00, 0.07) |
| 29-60d | 106 | 0.2% | 109 | 0.1% | \$772 (379, 1601) | 82 (50, 158) | 42 (33, 52) | 2.1 (1.1, 3.8) | 66.5% (45.6%, 79.7%) | 29 (15, 43) | 77534 (60561, 106648) | 0.33 (0.18, 0.48) | 0.24 (0.08, 0.49) | 0.04 (0.00, 0.13) |
| 61-90d | 39 | 0.1% | 39 | 0.0% | \$1108 (355, 1932) | 121 (86, 193) | 45 (38, 55) | 2.4 (0.9, 4.4) | 62.1% (48.1%, 78.4%) | 29 (14, 46) | 74235 (59743, 108513) | 0.29 (0.11, 0.42) | 0.27 (0.11, 0.52) | 0.03 (0.00, 0.11) |
| > 90d | 155 | 0.2% | 156 | 0.1% | \$1028 (436, 2235) | 253 (170, 329) | 48 (40, 53) | 2.5 (1.4, 4.4) | 66.8% (45.1%, 83.6%) | 29 (16, 45) | 77575 (56909, 114072) | 0.27 (0.09, 0.50) | 0.25 (0.09, 0.48) | 0.00 (0.00, 0.07) |

ERG®=Episode Risk Group, NHW=Non-Hispanic White, ADI=Area Deprivation Index, AGI=Adjusted Gross Income, HCP=Health Care Provider, DC=Doctor of Chiropractic, PT=Physical Therapist, LAc=Licensed Acupuncturist
Cells with red text denote that the effect of first line service timing on measured attributes was found not to be significantly different from that of No First Line or Not Provided reference - (Mann-Whitney U p > 0.001) Cells with black text denote that the effect of first line service timing on measured attributes was found to be significantly different from that of No First Line or Not Provided reference (ref) - (Mann-Whitney U p < 0.001)

**Figure 3.**
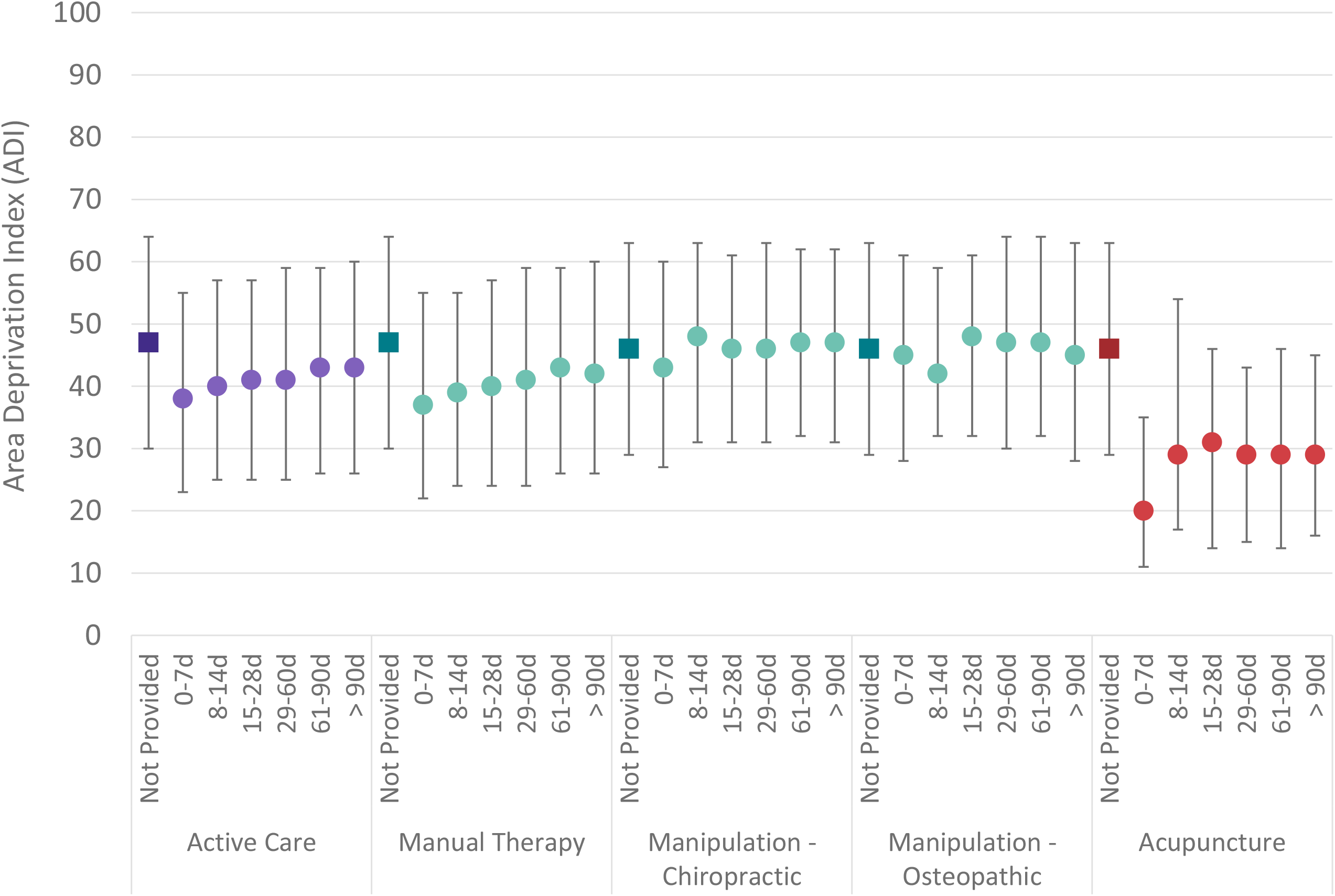
For individuals with neck pain initially contacting a PCP, Area Deprivation Index (ADI) of the individual’s home address zip code associated with median (Q1, Q3) number of days (d) into an episode when first line services are initially introduced

Compared to episodes without a non-pharmaceutical service, total episode cost was higher when any non-pharmaceutical service was provided at any time, except for OMT provided within 7 days of initial contact with a PCP. The total episode cost increase was lowest for introduction of CMT and OMT [Figure 4]. Episode duration increased as non-pharmaceutical services were introduced later in an episode [Table 3].

**Figure 4.**
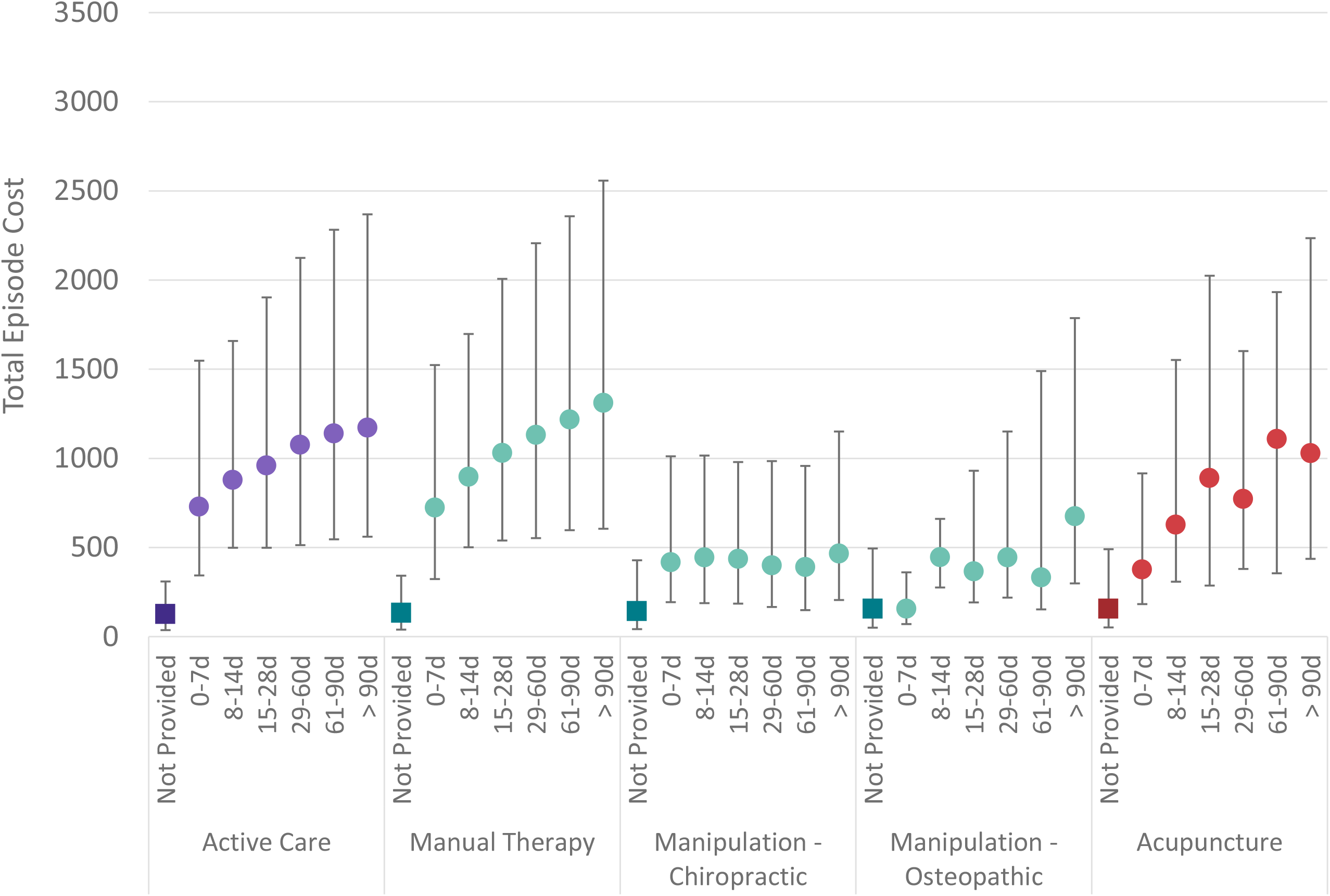
For individuals with neck pain initially contacting a PCP, median (Q1, Q3) total episode cost associated with number of days (d) into an episode when first line services are initially introduced

## Discussion

PCPs play an important role in the health care delivery system and are initially contacted by individuals with a variety of conditions, including NP. In the absence of serious pathology, NP CPGs generally recommend non-pharmaceutical and non-interventional approaches. Time burden, administrative complexity, cost, and other factors likely contribute to the observed low rate of use of CPG recommended non-pharmaceutical and non-interventional approaches for individuals with NP initially contacting a PCP. Strategies and tactics that make it easier for PCPs to provide CPG concordant care for NP is an important focus of translation efforts.

As a retrospective cohort study of associations, there are numerous potential limitations and confounders. The risk of selection bias was present due to the limited ability to control for individual preference and/or meaningful differences in clinical complexity of individuals seeking treatment of NP by a PCP versus other types of HCP, and individual expectations or requests for specific types of health care services. These limitations were partially addressed by limiting the cohort to only those individuals with non-surgical NP, and by excluding NP episodes associated with significant pathology.

Data errors, variability in benefit plan design, variability in enrollee cost-sharing responsibility, and missing information were potential sources of confounding or bias. These were partially addressed through limiting the cohort to only those having continuous highly uniform commercial insurance coverage and the processing of administrative claims data having included extensive quality and actuarial control measures. The identification of individual PCPs was based on data contained in a commercial insurer HCP database that may have included errors or missing information. Summarizing total episode cost has potential limitations associated with insurance coverage, nature of network participation, alternative reimbursement models, and individuals obtaining services outside of insurance coverage and reimbursement. The cohort, while including individuals from all 50 states and most US territories did not describe a U.S representative sample.

This study corroborates and expands on two earlier studies. First, an identical study of individuals with LBP initially contacting a PCP also found low rates of incorporation of AC, MT, CMT, OMT, and Acu.^47^ Second, a previous study found a low proportion of individuals with NP initially contacting a PCP have timely incorporation of guideline-concordant non-pharmacologic and non-interventional therapies.^35^ This study expands on this to explore more detailed distribution of episodes by timing of incorporation of AC, MT, CMT, OMT, and Acu. While these services are infrequently incorporated in NP episodes initially contacting a PCP, when incorporated AC, MT, CMT, OMT, and Acu tend to be introduced after 14 days into an episode. Future research should explore the degree to which patient self-selection versus PCP referral results in incorporation of AC, MT, CMT, OMT, and Acu for episodes of NP.

A previous study of spine-related disorders (SRDs), including NP, compared usual primary care, with a “primary spine care” (PSC) model.^58^ The PSC model was comprised of DCs, PTs and DOs embedded within a traditional primary care setting and directly accessed by individuals with a SRD. Compared to usual primary care, the PSC model was found to be associated with lower total costs in both the first and second year, with no meaningful differences in clinical outcomes. In this study, total costs were found to be higher when individuals with NP initially contacted a PCP and subsequently obtained treatment from a DC, DO, PT, or LAc. The absence of clinically meaningful differences in pharmaceutical, imaging, or interventional services indicates AC, MT, CMT, OMT, and Acu services were additive to typical primary care management. Further research comparing attributes of direct versus referral-based access to AC, MT, CMT, OMT, and Acu is warranted.

Previous studies of PCP referral patterns for LBP and chronic musculoskeletal pain found that administrative burden and the cost of non-pharmacologic therapies are perceived as barriers.^41,42^ This appears to be corroborated by this study’s finding that AC, MT, CMT, OMT, and Acu are infrequently incorporated by PCPs into management of NP, and if incorporated tend to be later in an episode after pharmaceutical, imaging, or interventional services. It was not possible to know whether AC, MT, CMT, OMT, and Acu were the result of PCP referral or individuals directly accessing these services after initially contacting a PCP.

This study of individuals with NP initially contacting a PCP is similar to previous LBP studies that found earlier use of non-pharmaceutical therapies may be associated with a reduction in use of low value services and prescription pharmaceuticals, including opioids. ^36,59,60^ The study finding that the benefits of early use of non-pharmaceutical therapies for NP are most evident if initiated within 7 days of initially contacting a PCP corroborates a similar finding for LBP ^61^, has potentially important translation implications, and warrants additional study given the relative absence of data for NP.

## Conclusion

A PCP is commonly the initial HCP consulted by individuals with NP or LBP. For individuals without red flags of serious pathology NP and LBP CPGs emphasize favorable natural history, self-care, and non-pharmaceutical services as first-line approaches. This study reveals that like LBP, individuals with NP initially contacting a PCP commonly receive pharmaceutical, imaging, and interventional services before non-pharmaceutical services. Non-pharmaceutical services are infrequently provided early in an episode when the potential benefits are greatest. Within the time constraints of a typical PCP visit, increasing early incorporation of guideline concordant non-pharmaceutical services likely involves making it easier for a PCP to address a variety of individual preferences, local socioeconomic, and HCP availability factors. As with LBP, making it easier for PCPs to suggest and individuals to follow through on recommendations to incorporate non-pharmaceutical services may be enhanced by a plain language summary of NP CPGs available to individuals before a visit with a PCP.

## Supporting information

Supplement - Risk Ratio

Supplement - State Summary

Supplement - STROBE Checklist

## Data Availability

All data produced in the present work are contained in the manuscript

## List of Abbreviations

NP: Neck pain
US: United States
CPG: Clinical practice guideline
PCP: Primary care provider
PS: Physician specialist
DC: Doctor of Chiropractic
PT: Physical Therapist
HCP: Health care provider I
QR: Interquartile range
OR: Odds ratio
RR: Risk ratio
AGI: Adjusted Gross Income
ADI: Area Deprivation Index
STROBE: Strengthening the Reporting of Observational Studies in Epidemiology
*CPT*^*®*^: Current Procedural Terminology
*ETG*^*®*^: Episode Treatment Group^*®*^
*ERG*^*®*^: Episode Risk Group^*®*^
ACP: American College of Physicians
PA: Physician Assistant
CMT: Chiropractic manipulative treatment
OMT: Osteopathic manipulative treatment
AC: Active care
MT: Manual therapy

## Notes

**Competing interests** At the time of manuscript submission **DE and MZ** are UnitedHealth Group employees and UNH stockholders. No other potential conflicts of interest or competing interests exist.

### Competing Interest Statement

The authors have declared no competing interest.

### Funding Statement

This study did not receive any funding

### Author Declarations

Because the data was de-identified or a Limited Data Set in compliance with the Health Insurance Portability and Accountability Act and customer requirements, the UnitedHealth Group Office of Human Research Affairs determined that this study was exempt from Institutional Review Board review.

## References

1. Safiri S, Kolahi AA, Hoy D, et al. Global, regional, and national burden of neck pain in the general population, 1990-2017: systematic analysis of the Global Burden of Disease Study 2017. Bmj. Mar 26 2020;368:m791. doi:10.1136/bmj.m791

2. Hurwitz EL, Randhawa K, Yu H, Cote P, Haldeman S. The Global Spine Care Initiative: a summary of the global burden of low back and neck pain studies. Eur Spine J. Sep 2018;27(Suppl 6):796–801. doi:10.1007/s00586-017-5432-9

3. Hoy D, March L, Woolf A, et al. The global burden of neck pain: estimates from the global burden of disease 2010 study. Ann Rheum Dis. Jul 2014;73(7):1309–15. doi:10.1136/annrheumdis-2013-204431

4. Ferrari R, Russell AS. Regional musculoskeletal conditions: neck pain. Best Pract Res Clin Rheumatol. Feb 2003;17(1):57–70. doi:10.1016/s1521-6942(02)00097-9

5. Diseases GBD, Injuries C. Global burden of 369 diseases and injuries in 204 countries and territories, 1990-2019: a systematic analysis for the Global Burden of Disease Study 2019. Lancet. Oct 17 2020;396(10258):1204–1222. doi:10.1016/S0140-6736(20)30925-9

6. Dionne CE, Rossignol M, Deyo RA, Koes B, Schoene M, Battie M. Back to the Future: A Report From the 16th International Forum for Back and Neck Pain Research in Primary Care and Updated Research Agenda. Spine (Phila Pa 1976). Oct 1 2022;47(19):E595–E605. doi:10.1097/BRS.0000000000004408

7. Dieleman JL, Cao J, Chapin A, et al. US Health Care Spending by Payer and Health Condition, 1996-2016. JAMA. Mar 3 2020;323(9):863–884. doi:10.1001/jama.2020.0734

8. Finley CR, Chan DS, Garrison S, et al. What are the most common conditions in primary care? Systematic review. 2018;64(11):832–840.

9. Collaborators UBoD. The State of US Health, 1990-2010: Burden of Diseases, Injuries, and Risk Factors. JAMA. 2013;310(6):591–606. doi:10.1001/jama.2013.13805

10. Childress MA, Stuek SJ. Neck Pain: Initial Evaluation and Management. Am Fam Physician. Aug 1 2020;102(3):150–156.

11. Elton D, Kosloff TM, Zhang M, et al. Low back pain care pathways and costs: association with the type of initial contact health care provider. A retrospective cohort study (preprint). medRxiv. 2022:2022.06.17.22276443. doi:10.1101/2022.06.17.22276443

12. Foster NE, Anema JR, Cherkin D, et al. Prevention and treatment of low back pain: evidence, challenges, and promising directions. Lancet. Jun 9 2018;391(10137):2368–2383. doi:10.1016/S0140-6736(18)30489-6

13. Meroni R, Piscitelli D, Ravasio C, et al. Evidence for managing chronic low back pain in primary care: a review of recommendations from high-quality clinical practice guidelines. Disabil Rehabil. Apr 2021;43(7):1029–1043. doi:10.1080/09638288.2019.1645888

14. Qaseem A, Wilt TJ, McLean RM, et al. Noninvasive Treatments for Acute, Subacute, and Chronic Low Back Pain: A Clinical Practice Guideline From the American College of Physicians. Ann Intern Med. Apr 4 2017;166(7):514–530. doi:10.7326/M16-2367

15. Parikh P, Santaguida P, Macdermid J, Gross A, Eshtiaghi A. Comparison of CPG’s for the diagnosis, prognosis and management of non-specific neck pain: a systematic review. BMC Musculoskelet Disord. Feb 14 2019;20(1):81. doi:10.1186/s12891-019-2441-3

16. Kjaer P, Kongsted A, Hartvigsen J, et al. National clinical guidelines for non-surgical treatment of patients with recent onset neck pain or cervical radiculopathy. Eur Spine J. Sep 2017;26(9):2242–2257. doi:10.1007/s00586-017-5121-8

17. Cote P, Wong JJ, Sutton D, et al. Management of neck pain and associated disorders: A clinical practice guideline from the Ontario Protocol for Traffic Injury Management (OPTIMa) Collaboration. Eur Spine J. Jul 2016;25(7):2000–22. doi:10.1007/s00586-016-4467-7

18. Corp N, Mansell G, Stynes S, et al. Evidence-based treatment recommendations for neck and low back pain across Europe: A systematic review of guidelines. Eur J Pain. Feb 2021;25(2):275–295. doi:10.1002/ejp.1679

19. Cohen SP, Hooten WM. Advances in the diagnosis and management of neck pain. BMJ. Aug 14 2017;358:j3221. doi:10.1136/bmj.j3221

20. Chou R, Cote P, Randhawa K, et al. The Global Spine Care Initiative: applying evidence-based guidelines on the non-invasive management of back and neck pain to low- and middle-income communities. Eur Spine J. Sep 2018;27(Suppl 6):851–860. doi:10.1007/s00586-017-5433-8

21. Bussieres AE, Stewart G, Al-Zoubi F, et al. The Treatment of Neck Pain-Associated Disorders and Whiplash-Associated Disorders: A Clinical Practice Guideline. J Manipulative Physiol Ther. Oct 2016;39(8):523–564 e27. doi:10.1016/j.jmpt.2016.08.007

22. Bryans R, Decina P, Descarreaux M, et al. Evidence-based guidelines for the chiropractic treatment of adults with neck pain. J Manipulative Physiol Ther. Jan 2014;37(1):42–63. doi:10.1016/j.jmpt.2013.08.010

23. Blanpied PR, Gross AR, Elliott JM, et al. Neck Pain: Revision 2017. J Orthop Sports Phys Ther. Jul 2017;47(7):A1–A83. doi:10.2519/jospt.2017.0302

24. Bier JD, Scholten-Peeters WGM, Staal JB, et al. Clinical Practice Guideline for Physical Therapy Assessment and Treatment in Patients With Nonspecific Neck Pain. Phys Ther. Mar 1 2018;98(3):162–171. doi:10.1093/ptj/pzx118

25. Beneciuk JM, Osborne R, Hagist MB, et al. American Physical Therapy Association Clinical Practice Guideline Implementation for Neck and Low Back Pain in Outpatient Physical Therapy: A Nonrandomized, Cross-sectional Stepped-Wedge Pilot Study. J Orthop Sports Phys Ther. Feb 2022;52(2):113–123. doi:10.2519/jospt.2022.10545

26. Neck Pain: Clinical Practice Guidelines Help Ensure Quality Care. J Orthop Sports Phys Ther. Jul 2017;47(7):513. doi:10.2519/jospt.2017.0508

27. Reid RO, Rabideau B, Sood N. Low-Value Health Care Services in a Commercially Insured Population. JAMA Intern Med. Oct 1 2016;176(10):1567–1571. doi:10.1001/jamainternmed.2016.5031

28. Schwartz AL, Landon BE, Elshaug AG, Chernew ME, McWilliams JM. Measuring low-value care in Medicare. JAMA Intern Med. Jul 2014;174(7):1067–76. doi:10.1001/jamainternmed.2014.1541

29. Østbye T, Yarnall KS, Krause KM, Pollak KI, Gradison M, Michener JL. Is there time for management of patients with chronic diseases in primary care? Ann Fam Med. May-Jun 2005;3(3):209–14. doi:10.1370/afm.310

30. Yarnall KS, Pollak KI, Østbye T, Krause KM, Michener JL. Primary care: is there enough time for prevention? Am J Public Health. Apr 2003;93(4):635–41. doi:10.2105/ajph.93.4.635

31. AlEissa SI, Tamai K, Konbaz F, et al. SPINE20 A global advocacy group promoting evidence-based spine care of value. Eur Spine J. Aug 2021;30(8):2091–2101. doi:10.1007/s00586-021-06890-5

32. Buchbinder R, Underwood M, Hartvigsen J, Maher CG. The Lancet Series call to action to reduce low value care for low back pain: an update. Pain. Sep 2020;161 Suppl 1:S57–S64. doi:10.1097/j.pain.0000000000001869

33. Cassel CK, Guest JA. Choosing wisely: helping physicians and patients make smart decisions about their care. JAMA. May 2 2012;307(17):1801–2. doi:10.1001/jama.2012.476

34. Colla CH, Morden NE, Sequist TD, Schpero WL, Rosenthal MB. Choosing wisely: prevalence and correlates of low-value health care services in the United States. J Gen Intern Med. Feb 2015;30(2):221–8. doi:10.1007/s11606-014-3070-z

35. Elton D, Zhang, Meng. Neck pain care pathways and costs: association with the type of initial contact health care provider. A retrospective cohort study (preprint). medRxiv. 2022;doi:10.1101/2022.07.18.22277777v1

36. Fritz JM, Kim M, Magel JS, Asche CV. Cost-Effectiveness of Primary Care Management With or Without Early Physical Therapy for Acute Low Back Pain: Economic Evaluation of a Randomized Clinical Trial. Spine (Phila Pa 1976). Mar 2017;42(5):285–290. doi:10.1097/BRS.0000000000001729

37. Kazis LE, Ameli O, Rothendler J, et al. Observational retrospective study of the association of initial healthcare provider for new-onset low back pain with early and long-term opioid use. BMJ Open. Sep 20 2019;9(9):e028633. doi:10.1136/bmjopen-2018-028633

38. Horn ME, George SZ, Fritz JM. Influence of Initial Provider on Health Care Utilization in Patients Seeking Care for Neck Pain. Mayo Clin Proc Innov Qual Outcomes. Dec 2017;1(3):226–233. doi:10.1016/j.mayocpiqo.2017.09.001

39. Zheng P, Kao MC, Karayannis NV, Smuck M. Stagnant Physical Therapy Referral Rates Alongside Rising Opioid Prescription Rates in Patients With Low Back Pain in the United States 1997-2010. Spine (Phila Pa 1976). May 1 2017;42(9):670–674. doi:10.1097/BRS.0000000000001875

40. Stevens GL. Behavioral and access barriers to seeking chiropractic care: a study of 3 New York clinics. J Manipulative Physiol Ther. Oct 2007;30(8):566–72. doi:10.1016/j.jmpt.2007.07.015

41. Roseen EJ, Conyers FG, Atlas SJ, Mehta DH. Initial Management of Acute and Chronic Low Back Pain: Responses from Brief Interviews of Primary Care Providers. J Altern Complement Med. Mar 2021;27(S1):S106–S114. doi:10.1089/acm.2020.0391

42. Penney LS, Ritenbaugh C, Elder C, Schneider J, Deyo RA, DeBar LL. Primary care physicians, acupuncture and chiropractic clinicians, and chronic pain patients: a qualitative analysis of communication and care coordination patterns. BMC Complement Altern Med. Jan 25 2016;16:30. doi:10.1186/s12906-016-1005-4

43. Greene BR, Smith M, Allareddy V, Haas M. Referral patterns and attitudes of primary care physicians towards chiropractors. BMC Complement Altern Med. Mar 1 2006;6:5. doi:10.1186/1472-6882-6-5

44. Coulter ID, Singh BB, Riley D, Der-Martirosian C. Interprofessional referral patterns in an integrated medical system. J Manipulative Physiol Ther. Mar-Apr 2005;28(3):170–4. doi:10.1016/j.jmpt.2005.02.016

45. George SZ, Lentz TA, Goertz CM. Back and neck pain: in support of routine delivery of non-pharmacologic treatments as a way to improve individual and population health. Transl Res. Aug 2021;234:129–140. doi:10.1016/j.trsl.2021.04.006

46. Dikkers MF, Westerman MJ, Rubinstein SM, van Tulder MW, Anema JR. Why Neck Pain Patients Are Not Referred to Manual Therapy: A Qualitative Study among Dutch Primary Care Stakeholders. PLoS One. 2016;11(6):e0157465. doi:10.1371/journal.pone.0157465

47. Elton D, Zhang M. Low back pain service utilization and costs: association with timing of first-line services for individuals initially contacting a primary care provider. A retrospective cohort study. medRxiv. 2022:2022.06.30.22277102. doi:10.1101/2022.06.30.22277102

48. Service IR. US Department of Treasury SOI tax stats - individual income tax statistics - 2017 ZIP code data. Accessed July 20, 2020. https://www.irs.gov/statistics/soi-tax-stats-individual-income-tax-statistics-2017-zip-code-data-soi.

49. Bureau USC. ACS Demographic and Housing Estimates. Accessed July 8, 2020. https://data.census.gov/cedsci/table?q=United%20States&g=0100000US&y=2018&tid=ACSDP1Y2018.DP05&hidePreview=true.

50. Kind AJH, Buckingham WR. Making Neighborhood-Disadvantage Metrics Accessible - The Neighborhood Atlas. N Engl J Med. Jun 28 2018;378(26):2456–2458. doi:10.1056/NEJMp1802313

51. von Elm E, Altman DG, Egger M, et al. The Strengthening the Reporting of Observational Studies in Epidemiology (STROBE) Statement: guidelines for reporting observational studies. Int J Surg. Dec 2014;12(12):1495–9. doi:10.1016/j.ijsu.2014.07.013

52. Hernan MA. The C-Word: Scientific Euphemisms Do Not Improve Causal Inference From Observational Data. Am J Public Health. May 2018;108(5):616–619. doi:10.2105/AJPH.2018.304337

53. Smith GD, Ebrahim S. Data dredging, bias, or confounding. BMJ. Dec 21 2002;325(7378):1437–8. doi:10.1136/bmj.325.7378.1437

54. King G NR. Why Propensity Scores Should Not Be Used for Matching. Political Analysis. 2019;27(4):435–454. doi:10.1017/pan.2019.11

55. National Academies of Sciences E, and Medicine. Health-Care Utilization as a Proxy in Disability Determination. 2 Factors That Affect Health-Care Utilization. National Academies Press 2018.

56. Insight O. Symmetry episode treatment groups: measuring health care with meaningful episodes of care. 2012.

57. Viera AJ. Odds ratios and risk ratios: what’s the difference and why does it matter? South Med J. Jul 2008;101(7):730–4. doi:10.1097/SMJ.0b013e31817a7ee4

58. Whedon JM, Toler AWJ, Bezdjian S, et al. Implementation of the Primary Spine Care Model in a Multi-Clinician Primary Care Setting: An Observational Cohort Study. J Manipulative Physiol Ther. Sep 2020;43(7):667–674. doi:10.1016/j.jmpt.2020.05.002

59. Fritz JM, Lane E, McFadden M, et al. Physical Therapy Referral From Primary Care for Acute Back Pain With Sciatica: A Randomized Controlled Trial. Ann Intern Med. Jan 2021;174(1):8–17. doi:10.7326/M20-4187

60. Fritz JM, Childs JD, Wainner RS, Flynn TW. Primary care referral of patients with low back pain to physical therapy: impact on future health care utilization and costs. Spine (Phila Pa 1976). Dec 1 2012;37(25):2114–21. doi:10.1097/BRS.0b013e31825d32f5

61. Liu X, Hanney WJ, Masaracchio M, et al. Immediate Physical Therapy Initiation in Patients With Acute Low Back Pain Is Associated With a Reduction in Downstream Health Care Utilization and Costs. Physical Therapy. 2018;98(5):336–347. doi:10.1093/ptj/pzy023

