## Supplement - Risk Ratio for "Neck pain service utilization and costs: association with timing of non-pharmaceutical services for individuals initially contacting a primary care provider. A retrospective cohort study"

**Supplement Risk Ratio - Individuals with non-surgical neck pain initially contacting a PCP - risk ratio and 95% confidence interval for service exposure based on timing of introduction of first line services compared to if service not introduced**

| | First Line | | | | | Passive Therapy | Second Line | | | | Third Line | | |
|---|---|---|---|---|---|---|---|---|---|---|---|---|---|
| | AC | MT | CMT | OMT | Acu | | Rx - NSAID | Rx - MM Relaxant | Imaging - Radiography | Imaging - MRI | Rx-Opioid | Spinal Injection | Imaging-CT |
| **Active Care (AC)** | | | | | | | | | | | | | |
| 0-7d | N/A | 35.56 (33.96, 37.24) | 3.54 (3.35, 3.73) | 1.84 (1.60, 2.11) | 3.30 (2.46, 4.44) | 12.17 (11.60, 12.76) | 0.65 (0.61, 0.68) | 0.66 (0.63, 0.70) | 1.39 (1.33, 1.45) | 1.27 (1.17, 1.39) | 0.63 (0.58, 0.69) | 4.36 (3.98, 4.77) | 0.53 (0.43, 0.65) |
| 8-14d | | 40.43 (38.54, 42.40) | 2.70 (2.47, 2.94) | 0.58 (0.41, 0.82) | 2.52 (1.57, 4.04) | 13.09 (12.35, 13.88) | 0.93 (0.87, 0.99) | 1.06 (1.01, 1.12) | 2.01 (1.92, 2.10) | 1.94 (1.76, 2.14) | 0.83 (0.75, 0.92) | 1.96 (1.63, 2.37) | 0.97 (0.78, 1.21) |
| 15-28d | | 39.25 (37.45, 41.14) | 2.85 (2.65, 3.08) | 0.60 (0.45, 0.82) | 3.64 (2.55, 5.19) | 12.71 (12.04, 13.42) | 1.07 (1.02, 1.13) | 1.10 (1.05, 1.15) | 2.05 (1.98, 2.13) | 2.88 (2.68, 3.09) | 0.95 (0.87, 1.03) | 2.21 (1.89, 2.59) | 1.23 (1.03, 1.46) |
| 29-60d | | 37.40 (35.66, 39.22) | 3.85 (3.62, 4.10) | 0.80 (0.62, 1.03) | 2.80 (1.89, 4.13) | 13.76 (13.08, 14.49) | 1.21 (1.15, 1.27) | 1.05 (1.00, 1.10) | 2.18 (2.10, 2.26) | 3.88 (3.66, 4.11) | 1.17 (1.09, 1.26) | 2.77 (2.42, 3.18) | 1.71 (1.48, 1.98) |
| 61-90d | | 35.17 (33.24, 37.21) | 5.23 (4.86, 5.63) | 0.93 (0.65, 1.33) | 4.76 (3.04, 7.45) | 14.93 (14.00, 15.93) | 1.44 (1.36, 1.53) | 1.05 (0.98, 1.12) | 2.08 (1.96, 2.19) | 4.28 (3.96, 4.63) | 1.35 (1.22, 1.50) | 3.25 (2.69, 3.91) | 1.86 (1.52, 2.28) |
| >90d | | 33.56 (31.95, 35.25) | 5.87 (5.60, 6.15) | 0.76 (0.59, 0.98) | 6.05 (4.62, 7.92) | 14.45 (13.76, 15.18) | 1.61 (1.55, 1.67) | 1.03 (0.98, 1.07) | 2.08 (2.00, 2.15) | 4.20 (3.98, 4.44) | 1.60 (1.51, 1.70) | 3.61 (3.21, 4.06) | 1.53 (1.32, 1.78) |
| **Manual Therapy (MT)** | | | | | | | | | | | | | |
| 0-7d | 16.56 (16.10, 17.04) | N/A | 2.41 (2.26, 2.58) | 0.64 (0.49, 0.82) | 6.23 (4.73, 8.20) | 10.05 (9.59, 10.53) | 0.66 (0.62, 0.70) | 0.70 (0.66, 0.74) | 1.20 (1.14, 1.27) | 1.22 (1.10, 1.35) | 0.65 (0.59, 0.71) | 3.32 (2.98, 3.70) | 0.43 (0.33, 0.56) |
| 8-14d | 18.21 (17.69, 18.73) | | 1.84 (1.65, 2.06) | 0.50 (0.33, 0.75) | 5.34 (3.56, 8.01) | 10.08 (9.49, 10.72) | 0.95 (0.88, 1.02) | 1.07 (1.01, 1.14) | 1.86 (1.77, 1.96) | 2.02 (1.81, 2.25) | 0.81 (0.72, 0.91) | 1.21 (0.94, 1.56) | 0.68 (0.51, 0.92) |
| 15-28d | 18.05 (17.56, 18.56) | | 2.07 (1.89, 2.27) | 0.54 (0.38, 0.76) | 5.87 (4.18, 8.24) | 10.27 (9.73, 10.83) | 1.03 (0.97, 1.09) | 1.15 (1.10, 1.21) | 1.96 (1.87, 2.04) | 2.86 (2.65, 3.08) | 0.93 (0.84, 1.02) | 1.89 (1.59, 2.26) | 1.29 (1.07, 1.55) |
| 29-60d | 17.70 (17.21, 18.21) | | 2.66 (2.47, 2.87) | 0.73 (0.54, 0.97) | 5.70 (4.08, 7.97) | 10.45 (9.93, 11.01) | 1.22 (1.16, 1.28) | 1.07 (1.02, 1.12) | 2.06 (1.98, 2.14) | 3.66 (3.44, 3.90) | 1.13 (1.04, 1.22) | 2.27 (1.94, 2.66) | 1.79 (1.53, 2.08) |
| 61-90d | 16.60 (16.01, 17.21) | | 3.64 (3.31, 3.99) | 0.92 (0.63, 1.36) | 7.01 (4.47, 10.98) | 11.10 (10.36, 11.90) | 1.37 (1.28, 1.47) | 1.10 (1.02, 1.18) | 2.06 (1.94, 2.18) | 4.37 (4.02, 4.75) | 1.37 (1.22, 1.53) | 2.71 (2.18, 3.36) | 1.77 (1.41, 2.24) |
| >90d | 16.19 (15.70, 16.69) | | 4.48 (4.25, 4.73) | 0.81 (0.62, 1.06) | 10.89 (8.47, 14.01) | 11.46 (10.92, 12.03) | 1.62 (1.56, 1.69) | 1.08 (1.03, 1.13) | 1.99 (1.92, 2.08) | 4.31 (4.08, 4.56) | 1.69 (1.59, 1.81) | 3.43 (3.02, 3.89) | 1.64 (1.40, 1.93) |
| **Manipulation - Chiropractic (CMT)** | | | | | | | | | | | | | |
| 0-7d | 3.88 (3.72, 4.06) | 2.85 (2.68, 3.04) | N/A | 0.43 (0.30, 0.61) | 5.95 (4.44, 7.99) | 9.36 (8.92, 9.83) | 0.67 (0.63, 0.72) | 0.78 (0.74, 0.83) | 1.32 (1.26, 1.40) | 0.70 (0.61, 0.81) | 0.77 (0.70, 0.85) | 3.89 (3.48, 4.35) | 0.54 (0.42, 0.70) |
| 8-14d | 3.32 (3.07, 3.58) | 2.70 (2.44, 2.99) | | 0.36 (0.19, 0.67) | 4.53 (2.71, 7.56) | 8.94 (8.31, 9.62) | 0.94 (0.86, 1.03) | 0.96 (0.89, 1.04) | 1.40 (1.29, 1.51) | 0.89 (0.73, 1.07) | 1.09 (0.96, 1.24) | 2.75 (2.23, 3.39) | 0.71 (0.49, 1.02) |
| 15-28d | 3.29 (3.08, 3.52) | 2.54 (2.32, 2.78) | | 0.31 (0.18, 0.55) | 5.89 (3.99, 8.68) | 8.97 (8.42, 9.56) | 0.97 (0.90, 1.04) | 0.83 (0.77, 0.89) | 1.30 (1.21, 1.40) | 0.89 (0.76, 1.05) | 1.14 (1.02, 1.26) | 2.03 (1.65, 2.51) | 0.67 (0.49, 0.92) |
| 29-60d | 3.34 (3.16, 3.53) | 2.70 (2.51, 2.90) | | 0.42 (0.28, 0.63) | 4.27 (2.91, 6.26) | 8.98 (8.50, 9.48) | 1.00 (0.94, 1.06) | 0.79 (0.74, 0.85) | 1.27 (1.19, 1.35) | 0.98 (0.86, 1.12) | 1.18 (1.08, 1.29) | 1.67 (1.38, 2.03) | 0.61 (0.46, 0.80) |
| 61-90d | 3.07 (2.85, 3.29) | 2.48 (2.26, 2.72) | | 0.58 (0.38, 0.88) | 5.07 (3.31, 7.78) | 8.75 (8.19, 9.34) | 1.21 (1.13, 1.29) | 0.90 (0.83, 0.97) | 1.27 (1.18, 1.37) | 1.00 (0.85, 1.17) | 1.39 (1.26, 1.53) | 1.55 (1.21, 1.98) | 0.67 (0.49, 0.93) |
| >90d | 3.45 (3.30, 3.61) | 2.84 (2.68, 3.01) | | 0.59 (0.44, 0.78) | 6.40 (4.89, 8.36) | 10.01 (9.58, 10.46) | 1.34 (1.28, 1.39) | 0.77 (0.73, 0.81) | 1.51 (1.45, 1.58) | 1.34 (1.22, 1.46) | 1.33 (1.24, 1.42) | 2.15 (1.87, 2.48) | 0.80 (0.66, 0.98) |
| **Manipulation - Osteopathic (OMT)** | | | | | | | | | | | | | |
| 0-7d | 0.88 (0.79, 0.98) | 0.49 (0.41, 0.58) | 0.42 (0.34, 0.51) | N/A | 2.06 (1.32, 3.21) | 1.00 (0.87, 1.14) | 0.28 (0.25, 0.32) | 0.36 (0.33, 0.40) | 0.34 (0.30, 0.38) | 0.35 (0.29, 0.44) | 0.29 (0.25, 0.35) | 1.05 (0.84, 1.31) | 0.25 (0.16, 0.38) |
| 8-14d | 1.08 (0.65, 1.79) | 0.83 (0.43, 1.61) | 0.12 (0.02, 0.84) | | 2.91 (0.41, 20.46) | 1.66 (0.98, 2.81) | 0.70 (0.47, 1.04) | 1.03 (0.78, 1.36) | 1.35 (1.01, 1.79) | 0.72 (0.33, 1.57) | 0.86 (0.51, 1.45) | 2.31 (1.07, 5.00) | 1.98 (0.91, 4.29) |
| 15-28d | 1.37 (0.96, 1.97) | 0.79 (0.45, 1.39) | 0.49 (0.22, 1.07) | | 4.00 (1.01, 15.86) | 1.62 (1.04, 2.52) | 0.83 (0.62, 1.12) | 1.18 (0.96, 1.44) | 0.93 (0.68, 1.26) | 0.50 (0.23, 1.09) | 1.03 (0.70, 1.53) | 1.85 (0.90, 3.81) | 1.13 (0.48, 2.68) |
| 29-60d | 1.67 (1.27, 2.20) | 1.78 (1.32, 2.41) | 0.78 (0.46, 1.31) | | 4.39 (1.42, 13.52) | 2.02 (1.45, 2.82) | 0.70 (0.53, 0.93) | 1.07 (0.88, 1.30) | 0.96 (0.74, 1.24) | 0.85 (0.51, 1.41) | 0.86 (0.60, 1.25) | 2.90 (1.79, 4.72) | 0.66 (0.25, 1.75) |
| 61-90d | 1.59 (1.05, 2.39) | 1.44 (0.87, 2.37) | 0.88 (0.43, 1.79) | | 3.08 (0.44, 21.68) | 2.35 (1.51, 3.65) | 1.17 (0.88, 1.56) | 1.27 (1.00, 1.61) | 0.65 (0.40, 1.04) | 1.54 (0.91, 2.59) | 1.29 (0.84, 1.97) | 3.27 (1.69, 6.32) | 3.15 (1.70, 5.84) |
| >90d | 1.90 (1.46, 2.47) | 2.15 (1.62, 2.85) | 1.19 (0.77, 1.84) | | 11.34 (5.47, 23.52) | 2.39 (1.74, 3.28) | 1.21 (0.99, 1.48) | 1.04 (0.85, 1.28) | 0.99 (0.76, 1.29) | 2.02 (1.46, 2.79) | 1.72 (1.33, 2.21) | 3.43 (2.15, 5.47) | 1.29 (0.62, 2.66) |
| **Acupuncture (Acu)** | | | | | | | | | | | | | |
| 0-7d | 1.66 (1.25, 2.21) | 3.31 (2.69, 4.07) | 2.22 (1.65, 3.00) | 4.05 (2.50, 6.58) | N/A | 4.59 (3.76, 5.61) | 0.34 (0.22, 0.53) | 0.31 (0.20, 0.49) | 0.64 (0.45, 0.90) | 0.59 (0.31, 1.12) | 0.62 (0.39, 0.99) | 2.69 (1.60, 4.54) | 0.18 (0.03, 1.26) |
| 8-14d | 2.30 (1.38, 3.85) | 3.80 (2.50, 5.78) | 2.01 (0.98, 4.14) | 1.39 (0.20, 9.54) | | 4.63 (2.96, 7.24) | 0.72 (0.37, 1.38) | 0.90 (0.54, 1.50) | 0.48 (0.19, 1.21) | 1.68 (0.75, 3.77) | 1.00 (0.45, 2.23) | 5.30 (2.37, 11.87) | 1.83 (0.48, 7.02) |
| 15-28d | 2.79 (2.01, 3.88) | 3.71 (2.70, 5.10) | 3.51 (2.43, 5.08) | 1.53 (0.39, 5.97) | | 4.26 (2.98, 6.07) | 0.85 (0.55, 1.31) | 0.74 (0.48, 1.15) | 0.87 (0.54, 1.40) | 2.23 (1.35, 3.69) | 1.32 (0.80, 2.19) | 5.27 (2.89, 9.61) | N/A |
| 29-60d | 2.70 (2.11, 3.46) | 4.55 (3.75, 5.52) | 4.03 (3.17, 5.14) | 2.85 (1.39, 5.83) | | 5.55 (4.50, 6.83) | 0.81 (0.59, 1.13) | 0.84 (0.63, 1.13) | 1.21 (0.91, 1.60) | 1.98 (1.33, 2.94) | 0.64 (0.37, 1.13) | 2.49 (1.28, 4.86) | 0.54 (0.14, 2.13) |
| 61-90d | 3.21 (2.25, 4.59) | 4.08 (2.85, 5.83) | 5.77 (4.32, 7.72) | 2.27 (0.59, 8.77) | | 6.33 (4.66, 8.60) | 1.35 (0.92, 1.96) | 0.59 (0.32, 1.09) | 0.99 (0.58, 1.69) | 3.04 (1.84, 5.02) | 1.80 (1.09, 2.97) | 5.22 (2.50, 10.91) | 1.50 (0.39, 5.80) |
| >90d | 3.92 (3.38, 4.54) | 5.40 (4.71, 6.18) | 5.71 (4.92, 6.62) | 1.99 (0.96, 4.11) | | 5.93 (5.04, 6.99) | 1.35 (1.11, 1.62) | 0.94 (0.75, 1.17) | 1.14 (0.90, 1.46) | 2.28 (1.68, 3.09) | 1.31 (0.96, 1.78) | 3.92 (2.53, 6.05) | 0.56 (0.18, 1.73) |

Confidence interval includes 1 for cells in red indicating risk is not different than the reference of if the specific service was not performed
