## Supplement - State Summary for "Neck pain service utilization and costs: association with timing of non-pharmaceutical services for individuals initially contacting a primary care provider. A retrospective cohort study"

| Supplement - Episode count by home address state of individual with neck pain | | | | | |
|---|---|---|---|---|---|
| **State** | **Episodes** | **%** | **State** | **Episodes** | **%** |
| **Total** | 137274 | 100.0% | **KY** | 1427 | 1.0% |
| **TX** | 16838 | 12.3% | **KS** | 1390 | 1.0% |
| **FL** | 13176 | 9.6% | **SC** | 1355 | 1.0% |
| **CA** | 6438 | 4.7% | **OR** | 1243 | 0.9% |
| **OH** | 6381 | 4.7% | **MA** | 1229 | 0.9% |
| **IL** | 6142 | 4.5% | **CT** | 1139 | 0.8% |
| **NC** | 6036 | 4.4% | **RI** | 1125 | 0.8% |
| **CO** | 5083 | 3.7% | **UT** | 984 | 0.7% |
| **AZ** | 5056 | 3.7% | **AL** | 857 | 0.6% |
| **WI** | 4957 | 3.6% | **NV** | 742 | 0.5% |
| **MO** | 4947 | 3.6% | **NM** | 534 | 0.4% |
| **MD** | 4227 | 3.1% | **DC** | 372 | 0.3% |
| **MN** | 4190 | 3.1% | **WV** | 288 | 0.2% |
| **GA** | 4044 | 3.0% | **ND** | 232 | 0.2% |
| **VA** | 3642 | 2.7% | **NH** | 227 | 0.2% |
| **IN** | 3332 | 2.4% | **ID** | 202 | 0.2% |
| **TN** | 3256 | 2.4% | **ME** | 167 | 0.1% |
| **NY** | 3153 | 2.3% | **DE** | 144 | 0.1% |
| **LA** | 3111 | 2.3% | **WY** | 124 | 0.1% |
| **NE** | 2256 | 1.6% | **SD** | 123 | 0.1% |
| **PA** | 2169 | 1.6% | **MT** | 77 | 0.1% |
| **AR** | 2114 | 1.5% | **VI** | 66 | 0.1% |
| **OK** | 2042 | 1.5% | **VT** | 46 | 0.0% |
| **WA** | 1980 | 1.4% | **PR** | 27 | 0.0% |
| **MS** | 1919 | 1.4% | **AK** | 24 | 0.0% |
| **IA** | 1902 | 1.4% | **HI** | 19 | 0.0% |
| **NJ** | 1827 | 1.3% | **Unknown** | 1156 | 0.8% |
| **MI** | 1737 | 1.3% | | | |
